# Super-Resolution of Magnetic Resonance Images Acquired Under Clinical Protocols using Deep Attention-based Method

**DOI:** 10.1101/2022.01.24.22269144

**Authors:** Bryan M. Li, Leonardo V. Castorina, Maria del C. Valdés-Hernández, Una Clancy, Stewart J. Wiseman, Eleni Sakka, Amos J. Storkey, Daniela Jaime Garcia, Yajun Cheng, Fergus Doubal, Michael T. Thrippleton, Michael Stringer, Joanna M. Wardlaw

## Abstract

Vast quantities of Magnetic Resonance Images (MRI) are routinely acquired in clinical practice but, to speed up acquisition, these scans are typically of a quality that is sufficient for clinical diagnosis but sub-optimal for large-scale precision medicine, computational diagnostics, and large-scale neuroimaging research. Here, we present a critic-guided framework to upsample low-resolution (often 2D) MRI scans. In addition, we incorporated feature-importance and self-attention methods into our model to improve the interpretability of this work. We evaluate our framework on paired low- and high-resolution brain MRI structural full scans (i.e. T1-, T2-weighted and FLAIR sequences are simultaneously input) obtained in clinical and research settings from scanners manufactured by Siemens, Phillips and GE. We showed that the upsampled MRIs are qualitatively faithful to the ground-truth high-quality scans (`PSNR` = 35.39; `MAE` = 3.78`E` −3; `NMSE` = 4.32`E` −10; `SSIM` = 0.9852; mean normal-appearing grey/white matter ratio intensity differences ranging from 0.0363 to 0.0784 for FLAIR, from 0.0010 to 0.0138 for T1-weighted and from 0.0156 to 0.074 for T2-weighted sequences). The automatic raw segmentations of tissues and lesions using the super-resolved images have fewer false positives and higher accuracy than those obtained from interpolated images in protocols represented with more than three sets in the training sample, making our approach a strong candidate for practical application in clinical research.

## 1 Introduction

Due to its non-invasive nature and radiation-free imaging approach, Magnetic Resonance Imaging (MRI) is commonly used in clinical diagnosis to visualise soft-tissue body structures, such as, in our case, the brain. However, there is an inherent trade-off between the spatial resolution of the images and the time required for acquiring them. In clinical settings, to save time, scans are often obtained at the minimal spatial resolution that allows the visual neuro-radiological assessment. It is common, for example, to ensure high spatial resolution in only one of the 3D planes (e.g. axial/horizontal), and acquire these images spaced 5 mm or more from each other, which results in poor spatial resolution in other views (e.g. sagittal or coronal/vertical planes). Also, sequence parameters and acquisition protocols differ between patients and hospitals, mainly owed to differences in scanner manufacturers and clinical manifestations. This all yields these images often impossible to process by standardised pipelines, rendering them inadequate for automated precision medicine approaches and large-scale research. Data-driven methods to upsample scans obtained in clinical settings that can preserve relevant clinical features without introducing visual artifacts, could drastically speed up the diagnosis process. Hence, there is a need to automate the scan processing including providing clinically accurate priors to support clinical decision-making, in addition to facilitating clinical research [60, 14].

With the recent advances in deep learning, artificial neural networks have achieved human-level performance across a range of tasks, such as image classification, natural language translation and protein structure predictions to name a few [23, 63, 28]. Because they are able to self-learn and extract features from high dimensional data, artificial neural networks have shown exceptional capabilities in increasing the spatial resolution (i.e. known as super-resolution (SR)) of single images [17, 33, 69]. Therefore, using artificial neural networks to increase the spatial resolution of clinically-acquired MRI holds potential and have seen rapid development in recent years [22, 46, 8]. If successful, this could be especially useful for potentially reducing acquisition time requirements or reconstruct poor-quality sequences within a scanning session. Retrospective assessment of low quality or low resolution data, for example, to investigate whether a pathology identified at a later point in time was already present (or not) in an earlier scan of the same patient, could also benefit from SR algorithms.

### 1.1 MRI Super-Resolution (SR)

Research in the MRI SR field has been helped by large-scale and publicly available datasets such as Human Connectome Project (HCP) [20] and fastMRI [66]. Nevertheless, the vast majority of these publicly available research datasets only contain high-resolution MRI scans generated in highly expensive research environments that do not reflect clinical practice. Thus, downsampling methods (i.e. subsequently represented abbreviated as *f*_downsample_) are needed to synthesise the corresponding low-resolution scans, in order to obtain low-resolution (X) and high-resolution (Y) paired samples [8] to train and test these algorithms. The deep learning models presented so far are trained on the generated low-resolution scans 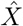 to learn to approximate the inverse function 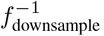, such that 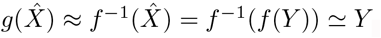. The caveat with this approach, is that the network is trained on “synthetic” low-resolution samples 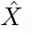, meaning that the output model may not perform as well on “real-world” low-resolution scans *X*, which may contain more artifacts than the estimated 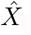.

Commonly used scaling methods for downsampling natural images such as nearest neighbour or bicubic interpolation, may not effectively represent the difference in quality between high- and low-resolution MRI scans [10]. K-space truncation has been used as an alternative for the production of low-resolution MR images [9]. These are obtained from applying the Fourier transform to the image obtained from the MRI scanner, where the intensity of each point represents the relative contribution of that point’s unique spatial frequency to the final image [6]. K-space truncation refers to the process of removing the points with higher frequencies, thus removing finer image details. Since MRI scans are obtained in k-space, this method is relatively straightforward for downsampling, but in addition to removing scan details, it can worsen visual artifacts such as Gibbs ringing, which appear as high frequency lines around areas of high contrast [6, 4, 5, 3]. A major problem of this downsampling method is that the low-resolution scans may contain consistent artifacts which can be falsely interpreted by deep learning methods as clues about the high-resolution scans, leading to poor performance on real-world data.

### 1.2 SR using Convolutional Neural Networks (CNNs)

Recently, SR methods have been implemented using deep learning techniques, facilitated by the growing capacity of computing power which allows the use of deeper architectures [46]. These are better than common linear interpolation methods [17], and relatively fast to compute due to the parallelization of calculations [35]. Several metrics and comparisons with conventional interpolation methods [7] show an indisputable advantage of these techniques in the problem at-hand. Convolutional layers are effective in extracting features from images and scans, and stacking multiple layers allow for a more complex representation of the extracted features [35].

The Super-Resolution Convolutional Neural Network (SRCNN) introduced by Dong et al. [17], was one of the first works that used convolutional neural networks (CNNs) to upsample RGB images. SRCNN first interpolates the low-resolution input image using bicubic interpolation to the same dimension of the target high-resolution image. The input image is, then, passed through two convolutional layers: the first extracts feature maps, while the second introduces non-linearity, mapping these features to a higher resolution representation. A final layer combines the predictions producing the reconstructed high-resolution image.This method outperformed those considered state-of-the-art at the time, such as Bicubic Interpolation, Sparse-Coding [64], Anchored Neighbourhood Regression [54], and Kernel Ridge Regression [31], in image upsampling tasks.

Dong et al. [18] further improved the SRCNN proposing the Fast SRCNN (FSRCNN). FSRCNN does not rely on the bicubic interpolation and, instead, uses three convolutional layers to extract the features, shrink the dimensionality to map the low-resolution features into the high-resolution feature space, and learn the non-linear mapping. Then, the dimensionalities are expanded and deconvolved into the output (i.e. reconstructed high-resolution image). This method could run 40 times faster than SRCNN while achieving similar or better results.

Building on their research, Kim et al. [30] proposed the use of Very Deep Residual Neural Networks (VDSR), which outperformed SRCNN in both, performance, as measured by Peak Signal to Noise Ratio (PSNR), and runtime, by the use of higher learning rates coupled with residual CNN layers to speed up the training process. Kim et al. [30] also made use of gradient clipping as a solution to the exploding gradients problem.

Shi et al. [51] used residual connection to enhance the SRCNN architecture with global residual learning and local residual learning, which aimed to overcome the partial loss of information that increases with the depth of neural networks. Their work outperformed popular architectures such as SRCNN, FSRCNN, and VDSR in terms of PSNR and Structural Similarity Index Measure (SSIM, Wang et al. 58). Pham et al. [46] applied similar techniques to MRI scans, using an architecture similar to SRCNN, but with residual skip connections between layers. They found that, using PSNR as their metric, residual architectures achieved a better performance compared to their non-residual counterpart, even when using a larger numbers of layers. Additionally, increasing the number of layers seemed beneficial only to residual architectures.

Although the above-mentioned convolution-based methods achieved remarkable results in image SR, they relied on conventional image metrics, such as mean-absolute error (MAE), PSNR and SSIM, to evaluate upsampled images against the target high-resolution images [17, 30, 51]. These metrics, albeit efficient to compute and easy to optimize, are not the most clinically relevant as they evaluate the quality of the entire image space (e.g. including background non-brain regions). In addition, both upsampled and target high-resolution images are needed in order to compute the mentioned image metrics, rendering it impossible to estimate how well the upsampled image is on inference.

### 1.3 SR using Generative Adversarial Networks (GANs)

Generative Adversarial Networks (GAN), first introduced in [21], are deep generative models that learn to generate compelling images which resemble the distributions of real datasets. GAN consist of two modules that “learn” back-to-back: a generator and a critic (also known as a discriminator). The generator learns to generate realistic images that mimic the true data distribution, whereas the critic learns to distinguish generated data from real data. This minimax optimization objective has seen tremendous success in data generation [29, 16, 34], unsupervised translation [70, 2] and many more [44, 62]. Subsequently, such critic-guided training objective has also been adopted into the realm of single-image SR [33, 57].

In the first SRGAN, in addition to the adversarial training, [33] implemented a perceptual loss function, which unlike pixel-based metrics like MAE and PSNR, calculates loss on relevant characteristics of the image. In this scheme, the total loss is calculated as the weighted sum between the adversarial loss and the results of this perceptual loss function (i.e. the VGG loss), which is a deep CNN per-se trained on the ImageNet dataset [27]. The VGG loss is calculated as the Euclidean distance between the reference image and a feature map from a pre-trained 19-layers VGG network truncated at the convolution, after activation and right before performing max pooling. The adversarial loss, obtained from the discriminator of the GAN applied to all training images, is calculated as the average classification accuracy of all training images (0 low-resolution, 1 high-resolution).

The SRGAN was later improved by truncating the perceptual VGG loss before the activation, which resulted in sharper edges, therefore being termed ESRGAN [57]. Subsequently, [36] used a two-stage SR Generator Network based on VDSR to generate a high-resolution image from which MSE loss is calculated. The pair of real and generated high-resolution images are also fed to a discriminator network which calculates a loss by distinguishing whether or not the input image is from the true or generated distributions. The pair also undergoes a Sobel filtering before calculating the edge loss. The study proposed an Edge Enhanced Hybrid (EEH) loss function which sums together the MSE loss, the Discriminator adversarial loss, and the edge loss [36]. This was proposed as an alternative to the perceptual loss, which, in the context of medical images such as MRI, tends to produce smoother images with spot artifacts, as the feature extraction of the VGG model is primarily trained on natural images. Additionally, training only on MSE loss usually results in blurred edges which the EEH loss avoids due to the Sobel Edge loss [36].

A potential disadvantage of GAN methods is that, although the generated output image has a seemingly “real” spatial intensity distribution, it may not match the input image [35]. To tackle this, You et al. [65] introduced GAN-CIRCLE, a framework based on CycleGAN [70], which consists of two GAN networks, with two generators and discriminators joined together. From an input image *x*, the forward GAN will produce a SR image *ŷ* which is then fed to the backward GAN to produce an estimate of the input image 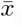, thus completing the circle. This circle is used to calculate the cycle-consistent loss 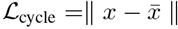 which aims at ensuring consistency between *x* and 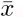. Additionally, other losses are taken into consideration into the overall objective function: 1) Adversarial loss from the discriminator, aimed at promoting consistent distributions in the output image; 2) Identity loss, fast to calculate, used for regularisation, and to refine the output image; 3) Joint Sparsifying Transform loss, calculated from two components: one which aims at maintaining general characteristics of the image such as anatomy, and a second one which reduces artifacts; and 4) Supervision loss, which is calculated from the paired real and generated samples P(*ŷ, y*) and 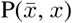. A weighted sum of these losses is, then, calculated.

### 1.4 Our Contributions

We use a dataset of paired low- and high-resolution scans of the same patients, who had an MRI head scan as part of a routine clinical examination, and, within 3 months, were re-scanned at a field strength of 3 Tesla using a research protocol, after consenting to participate on a study of sporadic small vessel disease (SVD) [11]. Thus, instead of using down-sampled scans 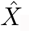, we use true low-resolution scans *X* acquired in routine clinical settings, for our framework to approximate the quality difference between low- and high-resolution scans directly *g*(*X*) ≈ *Y* independent from any choice of down-sampling function.

We developed an attention-based and critic-guided deep learning scheme that combines the advantages of both CNNs and GANs, and those of data fusion, to upsample clinical low-resolution whole MRI scans. Different from previous works that propose a SR algorithm to upsample a single MRI sequence despite referring to “MRI scans”, we propose and evaluate the use of data fusion for upsampling simultaneously the three MRI structural sequences most commonly acquired in a clinical MRI scan: T1-weighted, T2-weighted and fluid-attenuated inversion recovery (FLAIR). Our data fusion method considerably improves the quality of the upsampled full scans (i.e. of the three MRI sequences taken in the scan) both, qualitatively and quantitatively, through inter-channel pass-through of information.

We also explore the effect of 1) different image co-registration methods and 2) variations in the image acquisition parameters, in the performance of the proposed framework.

In addition, we explore the attention gates of the network to explain which details are extracted at each depth of its architecture, thus improving the explainability of the upsampling process. We also incorporated GradCAM [50] into the Critic to improve interpretability of the Critic model. Moreover, the learned activation maps can assist us in identifying potential artifacts in the upsampled images and not only upsample, but also reconstruct low-quality high-resolution scans.

To evaluate the applicability of our method in clinical research and, potentially, in clinical practice, 1) each of the test scans underwent the same image processing pipeline used in our clinical research studies to assess tissues and lesions, and 2) a neurologist and an experienced scan manager, independently and blind to each other’s results, visually assessed all the (full) scans involved in the test subsample, to evaluate the scope of the proposed framework and its feasibility for its further application to clinical research and practice.

## 2 Dataset

The data used in this study is part of an ongoing observational clinical study of sporadic SVD, a disease of the small blood vessels that, if progresses, can result in stroke and/or dementia, and is currently diagnosed by the presence of specific pathological features in brain MRI scans. The primary study that provided the data (Trial registration ISRCTN 12113543) includes a comprehensive set of assessments such as blood tests, blood pressure, retinal imaging, electrocardiograms, cognitive tests, and more to elucidate which factors determine the progression and rate of changes of the disease [11]. The present study only uses low-resolution and high-resolution paired brain MRI sets from 61 patients (mean age [std. dev.] 67.14 [9.78] years old, 19 females), each containing the structural MRI sequences T1-weighted, T2-weighted, and FLAIR. These sequences differently highlight tissues and neuroradiological features including pathology [47] and are acquired as part of all routine clinical MRI examinations. The only inclusion/exclusion criteria applied to select the sample used in this study was the acquisition of a paired low- and high-resolution scans within a small time-frame. The median time elapsed between both scans was 52 days (ranging from 8 to 95 days).

The low-resolution scans were obtained mainly at 1.5 Tesla (T) scanners from four different hospital sites in the Lothians and Fife regions in Scotland. Specifically, 34 were obtained from a 1.5T GE scanner model Signa-HDxt, 16 from a 1.5T Siemens scanner model Aera, 2 from a 1.5T Siemens Avanto, 1 from a 3T Siemens Prisma, 1 from a 1.5T Siemens Symphony Tim, 2 from a 3T Phillips Ingenia, 5 from a 1.5T Phillips Ingenia-Ambition X, and 1 from a 1.5T Phillips Ingenia. The sample comprises five different acquisition protocols in terms of spatial resolution and orientation of the three MRI sequences involved in the analyses. One example of a scan from each of these acquisition protocols included is shown in Figure 1, referred to subsequently as protocols 1 to 5. For sequences’ details, see Supplementary Material. The high-resolution scans were all obtained at 3T using a Siemens Prisma operating a research scanning protocol, at the Edinburgh Imaging Facility, using a 32-channel head coil, which facilitates high SNR and enables higher acceleration factors. Ethical approval for the primary study that provided the image data was obtained from South East Scotland Research Ethics Committee (Ref 18/SS/0044) on 31 May 2018, and from NHS Lothian Research & Development on 31 May 2018 (Ref 2018/0084).

**Figure 1:**
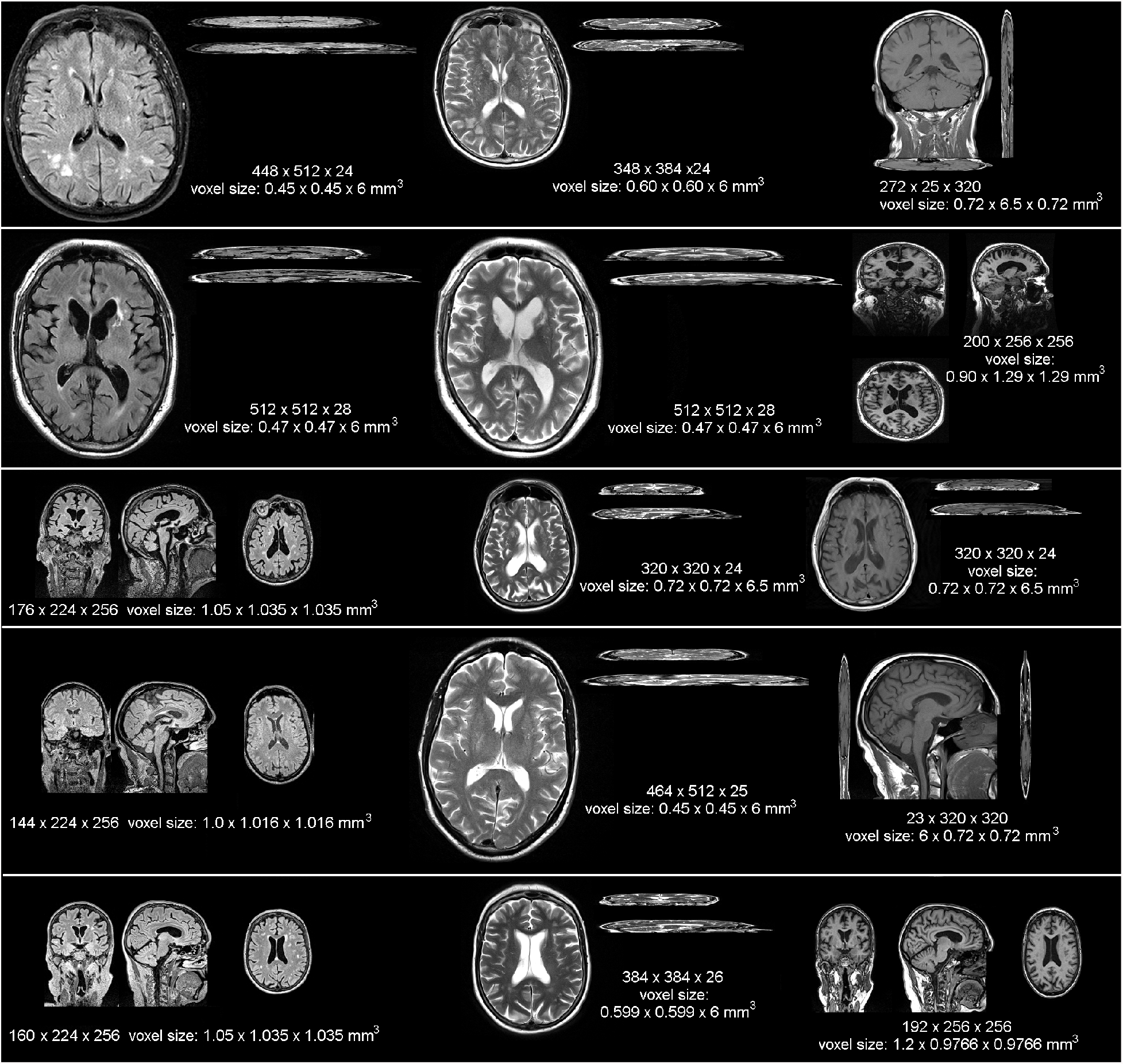
Samples images from each MRI scanner and acquisition protocol (in terms of spatial resolution and sequence orientation) that provided low resolution data for our analyses, displayed at the same scale to appreciate the heterogeneity in the native resolution and in terms of image contrast and orientation (i.e. 2D, 3D, sagittal, axial or coronal). From top to bottom, the protocols displayed are referred in the text as protocols 1 to 5.

The validation subsample included 10/61 patient datasets. In terms of the spatial resolution and orientation of the acquisition protocol of the low-resolution scans, 1/14 patient datasets of this subsample were from protocol 1, 6/36 from protocol 2, 2/6 from protocol 3, 0/1 from protocol 4, and 1/4 from protocol 5. Also, given the time elapsed between the high- and low-resolution scans, in our evaluation we used MRI data from the first 190 patients recruited for the same clinical study, acquired in subsequent visits separated up to three months (i.e. the time elapsed between the low- and high-resolution scans of our main sample), but all acquired with the high-resolution research protocol, as a “control” group (mean age [std. dev.] at recruitment 66.31 [11.22] years old, 59 females). Informed consent to participate in the parent study and for secondary use of anonymized data was obtained by all those who provided the images used in this study.

## 3 Methods

In this section, we detail our SR framework, including data preprocessing, generator and critic model architectures, evaluation metrics and inference procedure. An illustration of the complete workflow is shown in Figure 2.

**Figure 2:**
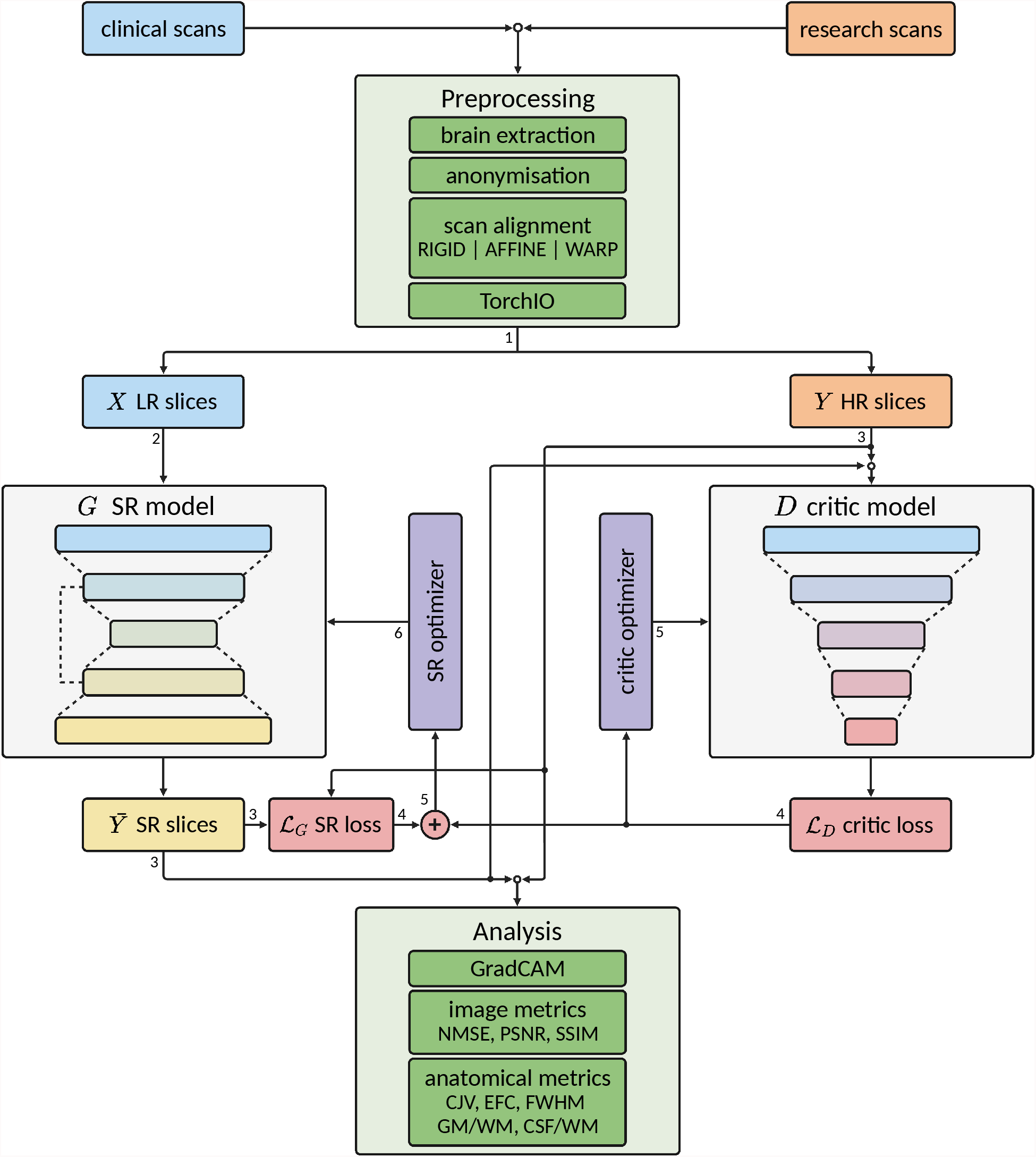
Illustration of the complete workflow of our super-resolution framework. Green coloured blocks indicate pre- and post-processing operations, grey coloured blocks indicate deep learning (DL) models, purple coloured blocks indicate optimizers for the DL models, light blue coloured blocks indicate low-resolution (LR) data, orange coloured blocks indicate high-resolution (HR) data, yellow coloured blocks indicate super-resolution (SR) data and red coloured blocks indicate loss calculations. Dark directed lines indicate the flow of the data and the number next to directed lines indicate the order of the data flow, note that some operations are performed concurrently.

### 3.1 Data Preprocessing

As part of the standard practice, an initial step of scanner magnet shimming is performed manually by the radiographers prior to acquiring any individual scan to reduce the b0 magnetic field inhomogeneities and optimise image quality, consequently influencing on the image intensities and contrast for displaying better the relevant clinical features.

Once the images are acquired and converted into NIfTI-1 format^2^, the preprocessing steps applied to all of them included correction of bias field inhomogeneities using FAST from the FMRIB Software Library (FSL) [61] to remove the slow-varying b1 magnetic field effects that could distort the images, and facilitate further brain extraction and scan alignment. The brain was extracted using the Brain Extraction Tool (BET2, Smith 52), also a tool from the FMRIB Software Library [61], to fully anonymize the data by removing all extracranial features (i.e. eyes, face, skin and skull) from the images. All intracranial volume binary masks obtained from this step were individually visually checked for accuracy and manually edited when/if required. The brain-extracted scan sequences were then aligned to an age-/atrophy-relevant brain template [26] in standard space, separately, in three ways:

- **Rigid**: using a rigid-body space transformation that involves only translations and rotations (i.e. 6 degrees of freedom). The shape and size (in volumetric units) of the individual brains are unaltered.
- **Affine**: using a linear transformation that, in addition to translations and rotations, also involves shearing and scaling (i.e. shrinking or enlarging, 12 degrees of freedom). This transformation, although preserves the shape by ensuring collinearity and preserving ratios of lengths of parallel planes’ segments, distorts the proportions of the brain structures/features among themselves: features located towards the cortex shrink or enlarge more than features at the centre of the brain.
- **Warp**: using a non-linear transformation aimed at achieving voxel-wise correspondence between the images involved, distorting not only the size but also the shape and morphology of the brain and all its features.

All space transformations were done with Niftyreg through TractoR [12] using the default parameters defined in Modat et al. [41]. However, as all registrations involved resampling and interpolation, to evaluate their effect against the results from our SR framework, linear space transformations were conducted in three ways: 1) using trilinear interpolation and the correlation ratio as cost function (default), 2) using sinc interpolation and mutual information as cost function, and 3) using spline interpolation and mutual information as cost function. After all images were aligned to the high resolution standard brain template, the intracranial volume of the template was used to crop the image space to reduce sparsity. The intensity values of the MRI scan sequences were linearly rescaled (i.e. linear normalisation) to the range [0, 1].

### 3.2 Network Input

TorchIO [45] was used to build the majority of the input pipeline. We represent each image *i* as a volume *V*_*i*_ with dimensions (*S, H, W*). In this study, each image has dimensions (148, 137, 135). We treat each volume as a 3D image of a brain slice (*S*), with height (*H*) and width (*W*). In order to upsample each slice using a 2D CNN, we expand the second dimension *C* in each image (i.e. *V*_*i*_ = (*S*, 1, *H, W*)), effectively, treating each slice as a gray-scale image. As each scan has 3 image sequences, namely FLAIR, T1-weighted and T2-weighted, and since each sequence captures different information, we combine all 3 sequences with the hypothesis that the data fusion of the three sequences can further improve the up-sampling performance [15]. To that end, we concatenate the sequences in the second dimension (i.e. (148, 3, 137, 135)) and feed the image with 3 sequences to the SR model.

We evaluate two different approaches of data input to the SR model:

- Slices are randomly selected, and entire slices with dimensions (*C, H, W*) are input to the model. Zero paddings are added to the spatial dimensions such that height and width are power-of-two values.
- Patches *B* with size (*P, P*) are randomly selected from each scan, each resulting in 3D input data of dimensions (*C, P, P*). This segmentation method is commonly used in other SR works in MRI [9, 38, 66]. Note that in this approach, depending on the number of patches *B*, not all regions of a scan may be exposed to the model.

We apply the data preparation procedure to both low-resolution and high-resolution scans in parallel to ensure the input data are in pairs. When training with mini-batching with batch size *N*, the size of the input to the model would be (*N, C, H, W*) or (*N, C, P, P*) depending on the preparation method, each requiring 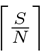 or 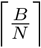 steps to train one scan respectively.

To explore the influence of training the model with FLAIR, T1-weighted and T2-weighted sequences simultaneously versus using each sequence separately, we treat each channel individually (i.e. (*N*, 1, *H, W*) or (*N*, 1, *P, P*)). The dataset was randomly split into train, validation, and test with a ratio of 66%-18%-16%, resulting in 40, 11 and 10 patient scans for each set respectively.

### 3.3 Model

The super-resolution model *G* used in this work is based on the U-Net architecture [49], a popular model commonly used in medical image segmentation tasks, which has also shown promising results in image super-resolution works [25, 39, 37]. Briefly, the U-Net architecture can be conceptually divided into two sub-modules. 1) encoder, where data features are compressed in the spatial dimension while increasing the number of channels, usually via consecutive blocks of convolutional and pooling layers. 2) decoder, which mirrors the encoder, though instead of compressing, it consists of transposed convolutional layers to decompress the data in its spatial dimension while decreasing the number of channels. The output of each convolutional block in the encoder is concatenated in the channel dimension with the inputs of the corresponding convolutional block in the decoder via residual connections. Such architecture have shown excellent results in extracting information from input features, and the addition of residual skip connections allow very deep networks to be used while mitigating the issue of vanishing or exploding gradients [24]. In our model, each convolutional block consists of a 2D convolutional layer with (3, 3) kernel and stride (1, 1), followed by a normalization layer and activation layer. We use Instance Normalization [55] and LeakyReLU (LReLU) activation [13]. The down-sampling block consists of a max pooling layer with (2, 2) kernel, which compresses the spatial dimension by a factor of 2, followed by a convolutional block. The up-sampling block consists of a transposed convolutional layer with stride size of (2, 2), to upsample the input variable by a factor of 2, followed by a convolutional block. When the shape of the residual connection and the input to the convolutional block differ, we apply zero padding to the variable with the smaller spatial dimension so that they can be concatenated. After the final convolutional layer, we scale the output logits to [0, 1] via a sigmoid layer to match the input data. Figure 3 illustrates the model architecture.

**Figure 3:**
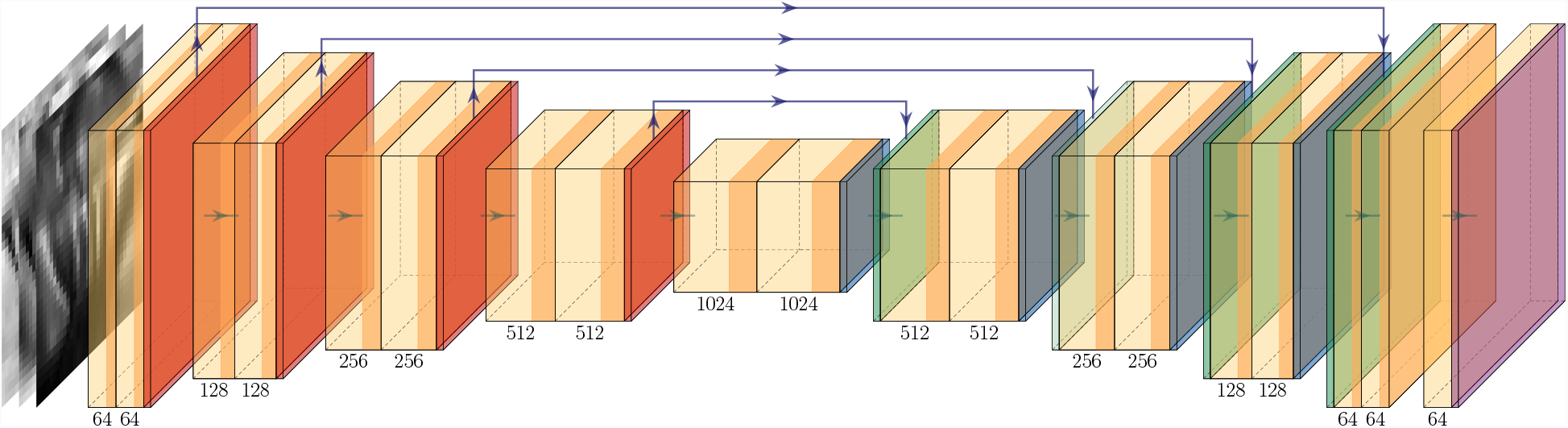
Up-sampling model *G* architecture. The two-shaded yellow block denotes a convolutional block, which consists of a convolutional layer followed by a normalization layer and activation layer. The orange, dark-blue, green and purple blocks denote down-sample, up-sample and output activation layer respectively. The green arrow between each block represents the flow of the data and the purple arrow above represents the residual connection. The number below each block indicates the number of filters used in the corresponding block.

### 3.4 Model Objective

To ensure that our framework can generalise on different datasets, we train the up-sampling model *G* using generic objective functions, including MAE, mean squared error (MSE), binary cross-entropy (BCE) and (maximize) SSIM, using Adam optimizer [32].

### 3.5 Critic

To overcome the issue of various image-based metrics not being able to capture the minute difference in MRI scans, we employ a separate critic network *D* to learn the distinction between high-resolution scan *y*_high_ and upsampled scan *ŷ*. The critic serves a very similar purpose as to the discriminator in a GAN, with the exception that our *G* and *D* networks can be trained separately.

The architecture of the critic model *D* follows a similar structure as the discriminator in DCGAN [48]. An illustration of the model is shown in Figure 4. The model consists of 4 consecutive blocks of convolutional layers with a kernel size of (4, 4) and stride size of (2, 2), each followed by an activation layer and dropout layer [53]. After the final dropout layer, a convolutional layer with 1 filter is used to compress the channel information to 1, and if the spatial dimension of the variable is larger than one, then an additional flatten layer is incorporated. The latent variable is then further compressed with a dense layer so that it returns a single scalar, which then followed by sigmoid activation to output a score value.

**Figure 4:**
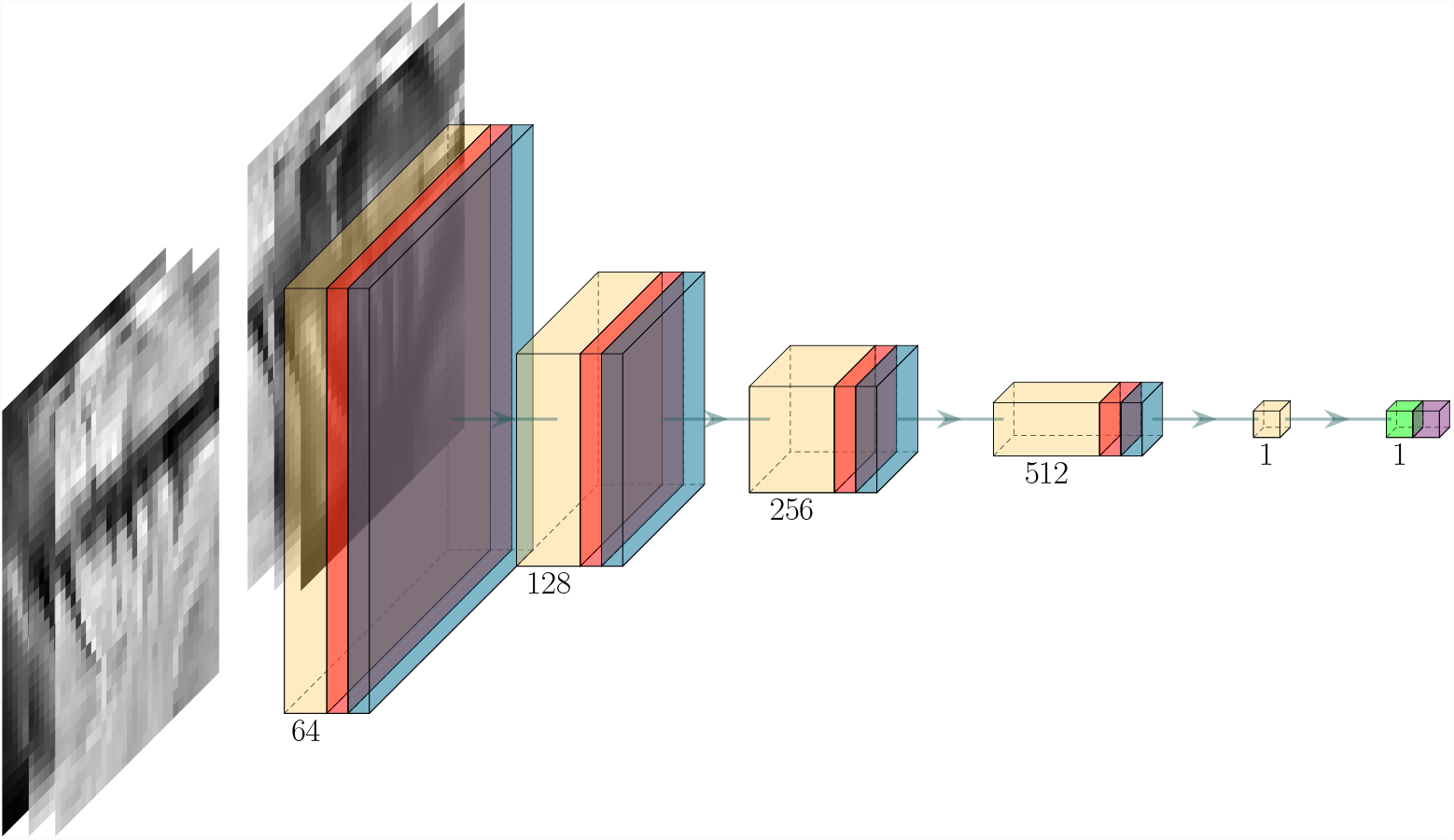
Critic architecture. Yellow, red, light blue, light green and purple block denote convolutional, activation, dropout, dense and output activation layer respectively. The green arrow between each block represents the flow of data and the number below each block indicates the number of filters used in the corresponding block.

Ideally, the critic should learn to predict a value of 1 when receiving a high-resolution input *y*_high_ and a value of 0 when receiving an upsampled input *ŷ*. We call this value the generation score ∼ [0, 1]. After each model step *ŷ*= *G*(*x*_low_), we feed both *ŷ* and *y*_high_ to the critic *D* and optimize the model with Adam optimizer at a learning rate of 2E-4 with the following loss function:

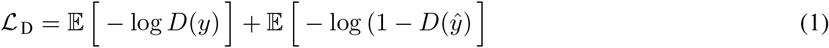

To ensure the critic can distinguish real scan against upsampled scans, we train *D* for multiple steps (i.e. 5 steps) for every *G* update. However, due to the very limited data, the critic model over-fits the training set very quickly, hence a high dropout rate of 0.5 is needed.

#### 3.5.1 Critic loss integration

In addition to use the critic generation score as a metric, we also use it to train our upsampling model *G*. We incorporated the generation score into our total upsampling model objective function as following:

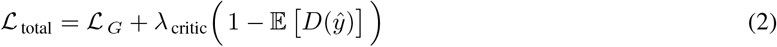

where the critic loss coefficient hyper-parameter *λ*_critic_ allow us to control the influence of the critic into the overall optimization objective.

### 3.6 Inference

The trained model can be used to upsample new data if the model is trained on slices (see section 3.2), as each slice of the MRI scan is passed to the model. However, we also randomly generated a number of patches for each scan to facilitate the training process with smaller GPU memory as has been proposed previously [38, 9, 27].

When patches were generated, each patch of a slice was upsampled. For reconstructing the output image, the upsampled patches were stitched together in a process that is handled by TorchIO [45]. The upsampled scans were then saved as a `.mat` file for further conversion to NIfTI-1 formatted files for clinical assessment by a medical professional.

### 3.7 Attention and Activation Maps

We Incorporated some of recently proposed methods to expand the explainability of our deep learning models. We adapted the Attention Gate module from Oktay et al. [42] into our up-sampling model *G*, in which the the model self-learns a set of masks to mask out regions in the inputs that are considered as less-relevant in the up-sampling task. We use four attention gates with dimensions 32, 64, 128, and 256. We stretch the learned masks (attention maps) and superimpose it onto the input image to obtain a heat-map of the attention at each point.

In addition to attention gated units, we analysed the activation map of the critic model using GradCAM [50]. Similarly, the activation map generated by GradCAM shows areas of relevance (or irrelevance) to the final predictions; though with one major distinction, the activation maps are not directly learn by the critic model.

### 3.8 Evaluation of the Results

#### 3.8.1 Image Quality Metrics

After reviewing the MRI SR literature from 2017 until May 2021 and compiling the list of metrics used by each of the papers included in Castorina et al. [7], we selected four of the metrics most frequently used to evaluate image quality: MAE, Normalized Mean Squared Error (NMSE), PSNR and SSIM, to evaluate the similarity between the upsampled and the “ground-truth” high-quality scan of our testing subsample, and, at the same time, allow comparability between our results and those already published, developed for the same purpose. Briefly, MAE is the absolute pixel-wise difference, averaged over the entire scan. NMSE is the ordinary MSE normalized by the value of the ground-truth pixel. The normalized squared error is given by

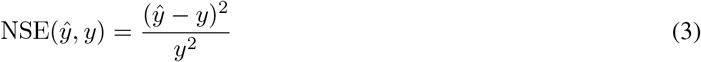

where *ŷ* and *y* are pixels at the same position of the upsampled and ground-truth scans, respectively. We compute the mean across all pixels from all image slices to obtain the NMSE. Similarly, PSNR captures the relation between the maximum value of a pixel and the MSE, and is given by

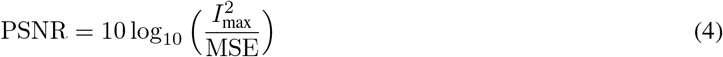

where *I*_max_ is the maximum value a pixel can take, and MSE is the unnormalized MSE. The SSIM [58] is a complex similarity metric that reflects the perceived similarity between two images more robustly than other individual metrics. For example, brightening a black-and-white image can lead to high NMSE and PSNR values yet the resulting image would be structurally identical to the original image and have a high SSIM.

While these metrics are useful in quantitatively assessing the overall quality of the upsampled images, they do not clearly reflect clinically meaningful results. We, therefore, in addition, computed the following metrics:

- **Coefficient of Joint Grey and White Matter Variation (CJV)**: Quantifies the intensity variability in white matter (WM) and grey matter (GM) accounting for the overlap between their distributions [19]. It is calculated as:

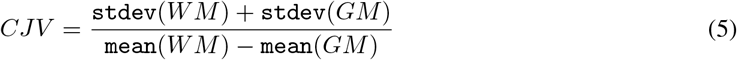
- **Entropy Focus Criterion (EFC)**: Uses the Shannon entropy of voxel intensities as an indication of ghosting and blurring induced by head motion [1].

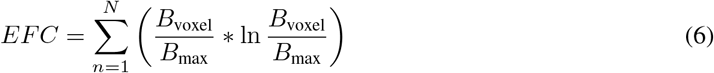

where *n* is the number of voxels and *B* is the voxel’s brightness.
- **Full-Width Half Maximum (FWHM)**: The full-width half maximum of the spatial distribution of the image intensity values in voxel units. Represents the smoothness of voxels.

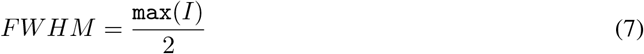

where *I* is the intensity across all voxels.
- **White-matter to maximum intensity ratio (WM2max)**: Median intensity within the WM mask over the 95% percentile of the full intensity distribution (i.e. calculated as the 95% confidence interval of the image intensities), with values around the interval [0.6, 0.8].
- **Ratio between the summary stats of grey matter (GM), white matter (WM) and cerebrospinal fluid (CSF)**: These are mean, standard deviation, 5% percentile and 95% percentile of the intensity distribution in CSF, GM and WM. For example:

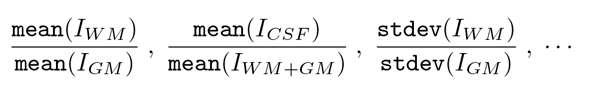

To ensure fair comparability, the GM, WM and CSF binary masks used in these evaluations were generated out from the brain-extracted T1-weighted images of the high-resolution subset using FSL-FAST [68]: a tool from the FMRIB Software Library.

#### 3.8.2 Applicability to Clinical Research - Structural image segmentation

In our testing sample, we automatically segmented lesions, normal tissues and anatomical structures in 1) images upsampled using our framework, 2) images upsampled from different types of interpolations (see Section 3.1), and 3) high-resolution images. We also segmented the same features in the images of the larger “control” sample, fully automatically, individually at each time point following the same procedure, to explore the variability in the longitudinal measurements using only high-resolution images acquired within the time frame equivalent to the time elapsed between the low- and high-resolution paired scans. CSF, veins and sinuses, pial layers and normal brain tissue were segmented using Gaussian clustering of a multidimensional array formed by combining the FLAIR, T2-weighted and T1-weighted images (i.e. upsampled or high-resolution). The multispectral clustering results were subsequently optimised using the expectation-maximization algorithm. CSF-filled spaces are identified as hypointense in T1-weighted and FLAIR, and hyperintense in T2-weighted. Venous sinuses and main venous pathways are hypointense in T2-weighted and with mid-to-low range intensities in T1-weighted and FLAIR. Pial layers, CSF-brain parenchyma partial volume effects and interstitial fluids, indistinguishable from each other and therefore classed together, are hypointense in T1-weighted and hyperintense in FLAIR and T2-weighted. Mid-range intensities in the three sequences correspond to normal-appearing grey and white matter tissues, each with characteristic contrast depending on the sequence.

Binary masks of ischaemic lesions (i.e. these including white matter hyperintensities (WMH) and stroke lesions) were generated from thresholding the intensity values of the brain-extracted FLAIR, with a threshold equal to 1.3*standard deviation above the mean, using an adjusted method from Zhan et al. [67]. Resulting hyperintense areas unlikely to reflect pathology were removed automatically using a lesion distribution template generated from the segmentation results of a large study of cognitive ageing [59]. Further refinement of the lesion segmentation was achieved by applying a Gaussian smoothing, followed by the removal of voxels with intensity values below 0.1 and with z-scores below 0.95. The WMH binary mask obtained from the high-resolution image was visually checked and manually edited to correct for inaccuracies and inclusion of the stroke lesion, for evaluating the raw results from the different upsampling methods, against the edited mask. The manual editing was performed using MRIcron^3^.

#### 3.8.3 Visual Assessment of Image Quality

A neurologist and an experienced scan manager, independently and blind to each other’s results, rated the quality of the scans of our test subsample: low-resolution, high-resolution as well as the upsampled images from the models that provided the best results using our SR framework, on a quality scale ranging from 0 to 5, being 5 high quality. Each rater was asked to outline the own criteria followed for rating the scans.

### 3.9 Code Availability

The codebase^4^ was predominantly written in Python 3.8, where the core part of the deep neural networks and model training procedures were implemented using the PyTorch [43] library and the scikit-image [56] library was used for image manipulation. We experimented with the model training and inferencing on various GPUs, including the NVIDIA RTX 2060, NVIDIA RTX 2080 Ti and NVIDIA V100, to ensure our model works with different levels of computing hardware. To further improve the model training performance, we have incorporated mixed-precision training where the majority of the computation was done in `float16` (half-precision) instead of `float32` [40], effectively allowing us to fit twice as much data onto the GPU memory. In addition to the implementation of our methods, the repository also contains clean, modular code for training, evaluating and applying models to different datasets with extensive logging functionality with TensorBoard^5^.

## 4 Results

In this section, we first, present the results from sampling random patches of the scans (Section 4.1) and full scan slices (Section 4.2) in a framework that used a common objective function as a loss function. In Section 4.3, we present the results from using full scan slices as input but in a framework trained with an adversarial loss instead. We present the results of evaluating the influence of the scan alignment techniques (i.e. rigid vs. affine vs. warping) in Section 4.4 and Section 4.5. Results of the implementation of attention and activation gates are presented in Section 4.6 and Section 4.7 respectively. We show the results from evaluating the applicability of our framework in clinical research and its potential for being introduced in clinical practice, in Section 4.8.

### 4.1 Upsampling Random Patches

Due to the large size of the MRI scans, sampling patches (i.e. as opposed to sampling whole slices) may be necessary, depending on available GPU memory. We evaluated the effects that: 1) variations in the content of the input data (i.e. multi-channel vs. single-channel), and 2) variations in key hyper-parameters, had on our three quantitative performance metrics.

#### 4.1.1 Multi-channel Versus Single Channel Input

We evaluated the effect of using a single MRI sequence as input, therefore processing the three sequences separately, versus using as input the three sequences (i.e. FLAIR, T1-weighted, T2-weighted) simultaneously (i.e. multi-channel input). As Figures A.1 and A.4 show, the models trained with the three scan sequences simultaneously input (i.e. data fusion) yielded significantly better performance. By using a multi-channel input data, information from different channels complement information that may have been missed in some areas. For instance, in Figure 5, the supramarginal gyri (highlighted in red), appear missing in the original low resolution FLAIR (i.e. due to interpolating thicker slices), but are present in the T1-weighted sequence and partially visible in T2-weighted. The upsampled scan from a model that uses the multi-channel input has the previously missing information present in all sequences, being the result qualitatively closer to the target high-resolution.

**Figure 5:**
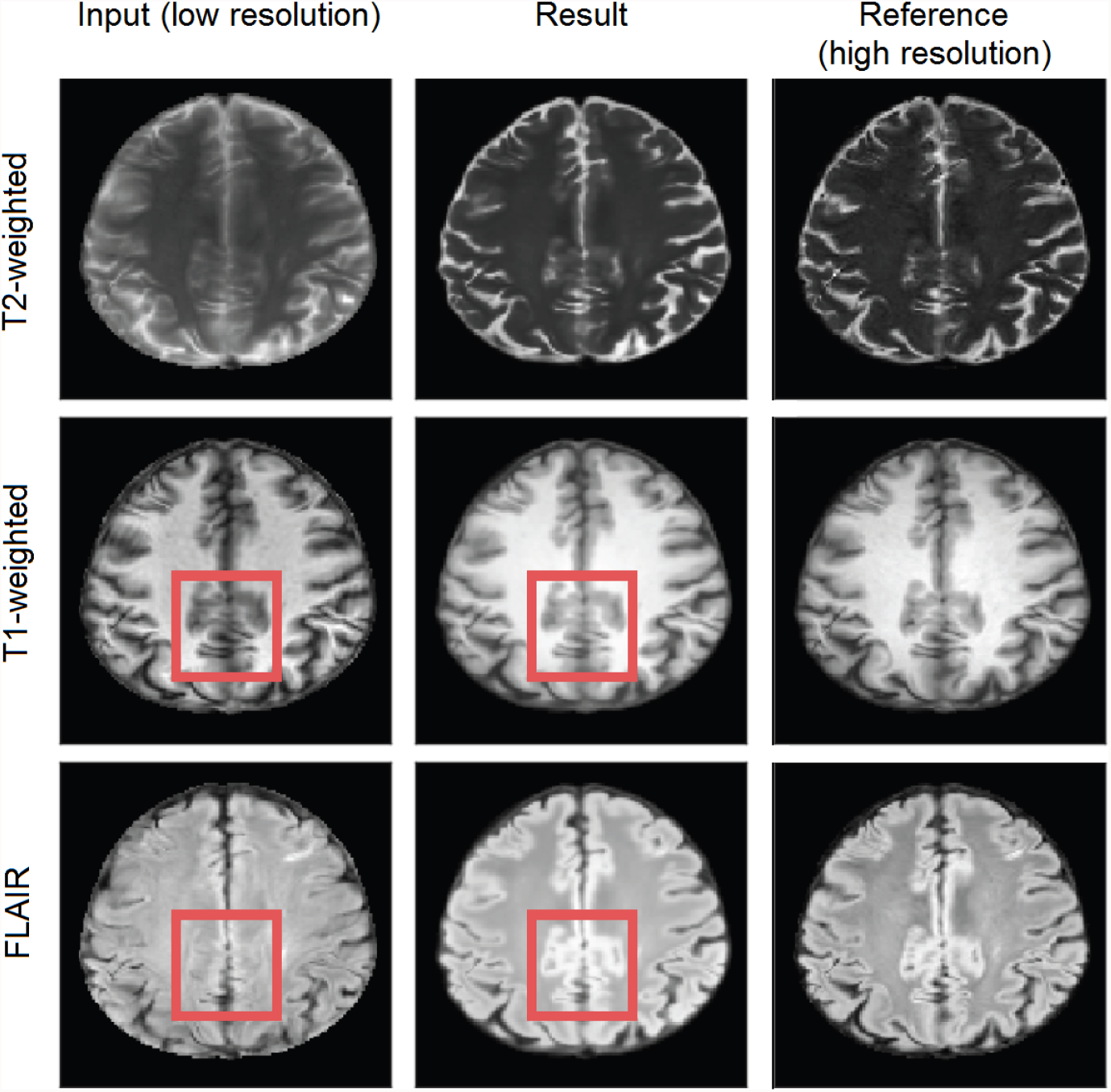
Left Column is the input image, the middle on is the generated (output) image and the right one is the target image. The red square represents an area of possible inter-channel pass-through information.

#### 4.1.2 Effect of Key Hyper-parameters - Patch Dimensions

We tested patches of dimensions: 8 × 8, 16 × 16, 32 × 32, 48× 48, and 64 × 64. Generally, the larger the patch size, the better the performance of all the three metrics (i.e. PSNR, SSIM and MAE). Larger patch sizes encompassed an increase in computational cost for training the model, as the batch size for larger patches is smaller and, therefore, training (and upsampling) takes longer. See Figure A.1 for results.

#### 4.1.3 Effect of Key Hyper-parameters - Number of Patches

The number of patches obtained from one scan influences the MRI space explored by the network. We evaluated our framework using 100, 500, 1000 and 5000 patches, given that the volume size of each sequence is (148, 137, 135). The more patches sampled, the higher was the performance. Extracting more patches came at a computational cost during training, but it generally resulted in higher upsampling (i.e. SR) performance. Additionally, as the larger the patch size the better the performance, the best performance was obtained when using a patch dimension of 64×64 with the largest number of patches evaluated (i.e. 5000 patches per scan) (see Figure A.2 and A.3).

#### 4.1.4 Effect of Key Hyper-parameters - Number of Filters

As shown in Figure A.4, generally, increasing the number of filters helped to improve the performance up to a certain point. Using 128 filters often yielded good performance but also showed signs of over-fitting. For our dataset, we found 64 filters to yield the best upsampling (i.e. SR) performance with the least computational requirements, offering a very good trade-off between performance, generalizability and expressiveness.

### 4.2 Upsampling Full Scans Slices

We evaluated using each scans’ slice as input versus patching, while using as input each sequence (i.e. FLAIR, T1-weighted and T2-weighted) simultaneously in a multi-channel approach versus treating each sequence separately.

#### 4.2.1 Patching Versus Full Slices

SR of full scan slices was possible although this required larger GPU memory. As shown in Figure A.4, using whole slices as input had a major increase in performance for single sequence SR (i.e. input data split per sequence). In the models that used multi-channel input (i.e. sequences merged in a single data array), training using whole slices as input produced only a slight improvement over patching but at an increased computational cost.

### 4.3 Upsampling Full Scans Slices with Adversarial Loss

The metrics commonly used for measuring the performance of SR models [7] at times fail to consider the upsampled scan as a whole and reflect just pixel-wise differences. We, therefore, implemented the use of a critic in the training process to distinguish between generated upsampled images and real high-resolution images. In this framework design, we calculate the error rate, which is then optionally added onto the loss function. As shown in the Supplementary Table A.1, adding the critic loss to the loss function did not seem to improve the performance. However, in our proposed framework, we still train the critic in conjunction with the conventional model (i.e. that uses MAE, SSIM and PSNR in the loss function) without adding on the loss, to use GradCAM to identify potential artifacts in the upsampled scan, as discussed in Section 4.7.

### 4.4 Comparing Scan Alignment Techniques

The numeric results obtained from using affine and rigid registration techniques for aligning the MRI sequences of the low resolution and high resolution scans to the high resolution standard brain template, can be seen in Table A.1. Warping consistently under-performed the linear registration techniques in all tests and did not render clinically useful results. From the two linear registration methods, the affine transformation gave slightly better results.

### 4.5 Best Model

As Table A.1 shows, the best model was Model 0, which used affine transformation to align the scans to the template, and full slices of the three MRI sequences (i.e. FLAIR, T1-weighted, and T2-weighted) input simultaneously. In our validation set we obtained `MAE` = 3.783`E` −03, `NMSE` = 4.32`E` −10, `PSNR` = 35.39, `SSIM` = 0.9852. In Figure 6 we present the upsampled scan for a patient with images acquired with protocol 2. The visual improvements are especially visible for FLAIR- and T2-weighted sequences, with higher contrast between white and grey matter, added details, and sharper (i.e. less blurry) images.

**Figure 6:**
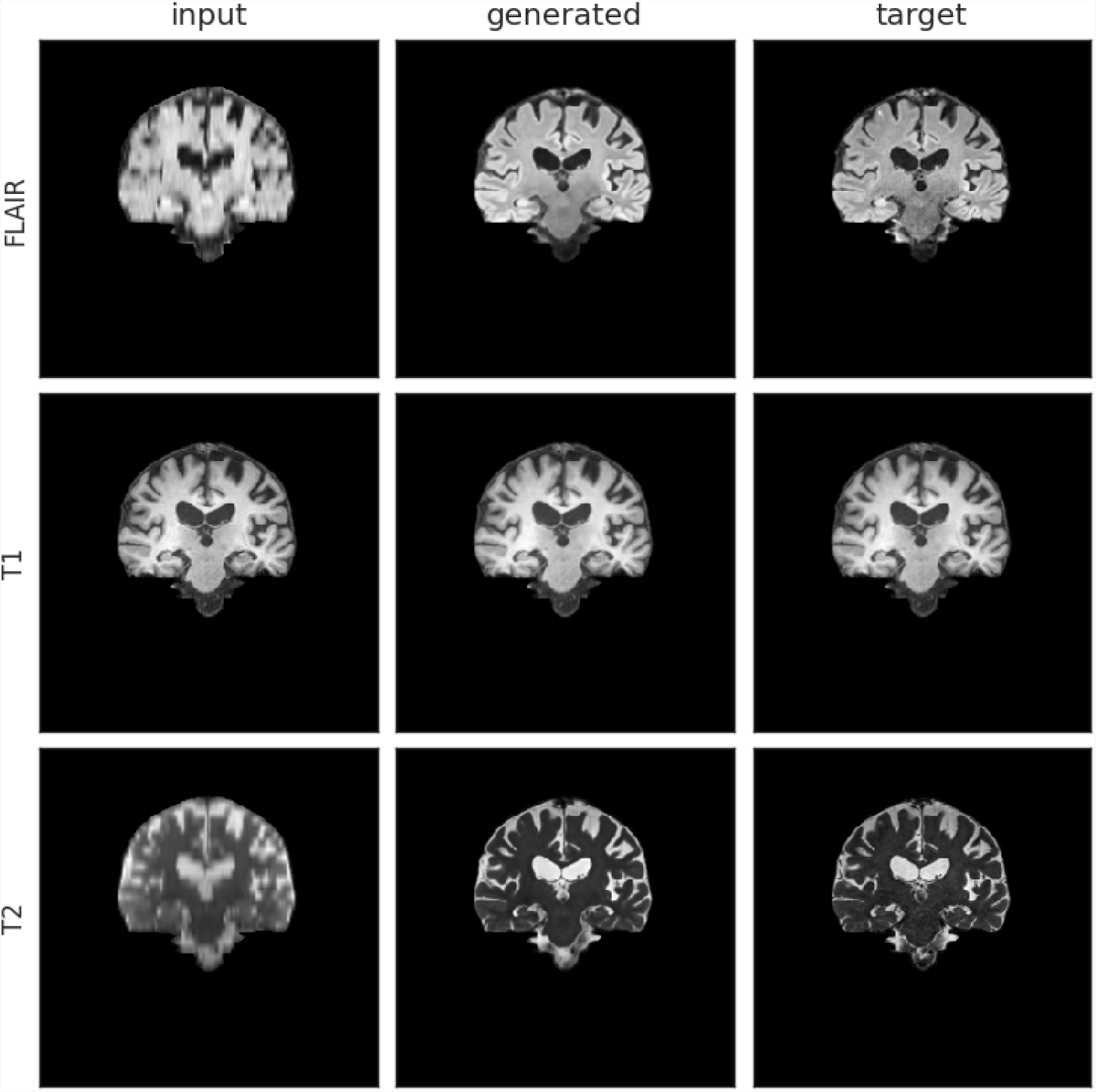
Comparison of generated upsampled and target scans for a patient with images acquired with protocol labelled as 2, produced by Model 0. Left Column shows the input images, the middle column shows the generated (output) images and the right column shows the target images.

From the models that used rigid-body transformation to align the input and target (i.e. reference) images to the template, Model 6 (which also used a multi-channeled full slice input approach) was the best. In the validation set we obtained `MAE` = 3.88`E` − 03, `NMSE` = 4.35`E` − 10, `PSNR` = 35.2, `SSIM` = 0.984 using this model.

We further compared the generated images of each model with the target high-resolution images plotting the difference maps as shown in Figure 7. As as per the example shown in the figure, the difference between the brain tissue of the target and the generated SR image for the T2-weighted sequence was very low across the sample, because the tissue contrast of this sequence in the native low-resolution and high-resolution images of all the data used in testing and training was similar. Large variations in T1-weighted and FLAIR sequence acquisition parameters across the dataset used caused differences between the upsampled and target image contrasts in these two sequences.

**Figure 7:**
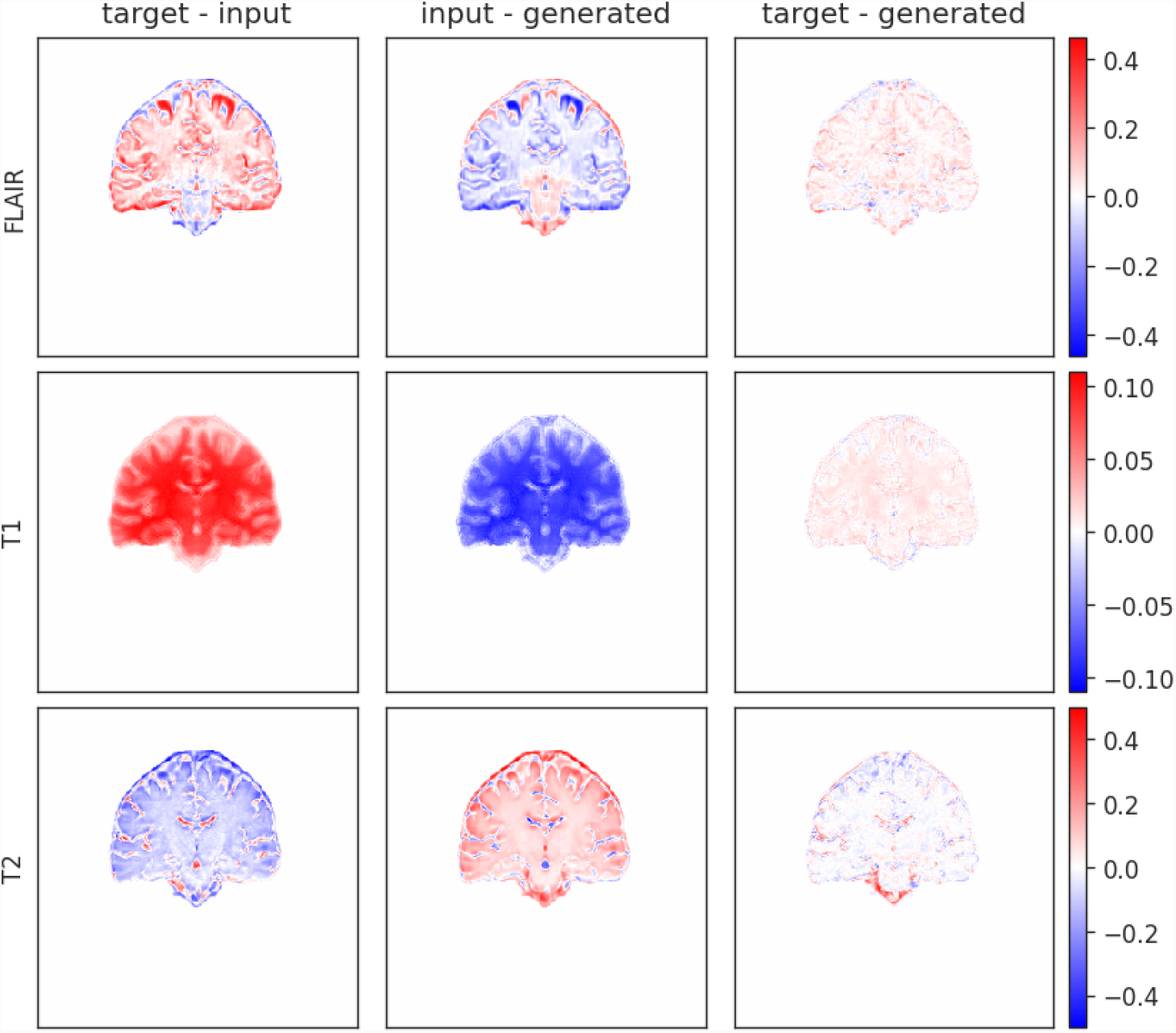
Difference map of input, target and generated upsampled scan from Model 0 for an image dataset from the acquisition protocol labelled as 2. The left column is the difference between target and input, the middle is between the input and the generated and the right is between target and generated.

### 4.6 Attention Gates Visualisation

The self-learned attention maps (see Section 3.7) in Figure 8 illustrate the areas of focus learned by each Attention Module in Model 0. This makes it possible to understand which areas of the convolved image are important for generating the SR image in the model that exhibited best performance. The green areas dominating the background across all gates indicate stable moderate attention.

**Figure 8:**
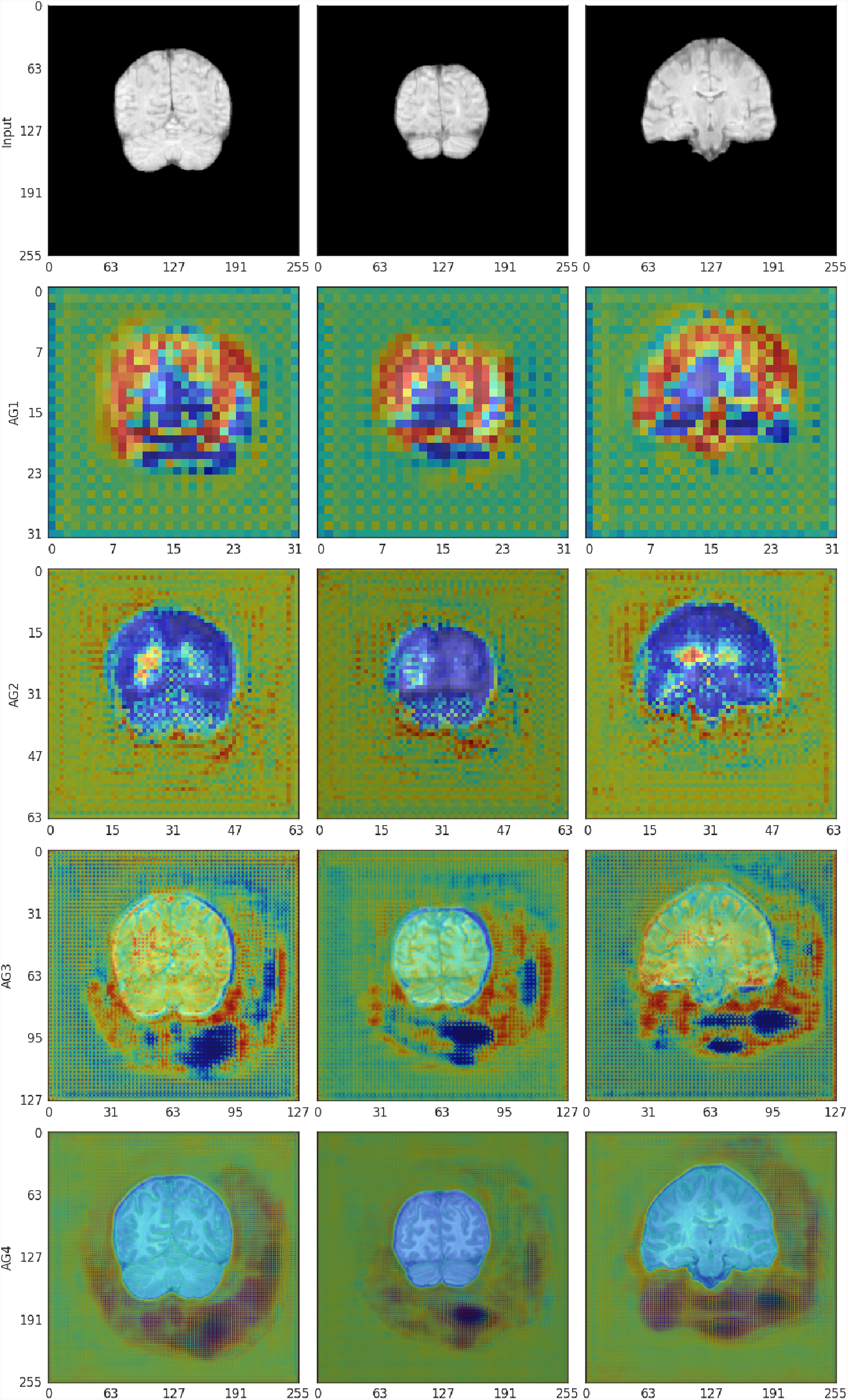
Attention Gates 1-4 from the Attention-Gated U-Net Upsampler for affine-registered scans (Model 0). Attention is represented in a `jet` colormap, with areas of high attention are in red and low attention in blue.

**Gate 1** (size 32) has moderate attention in the background and higher attention around the edges of the brain. Some areas of low attention are present inside the brain and at the cerebellum.

**Gate 2** (size 64) also has a moderate attention level in the background and lower in the brain tissue, especially in the white matter. Higher attention areas are located bordering the brain and around the brain ventricles.

**Gate 3** (size 128) has moderate attention in both the background and the brain tissue. Brain borders and sulci have contrasting low and high attention. Background areas bordering the brain with high and low attention in Gate 2 appear consolidated and extended in this gate.

**Gate 4** (size 256) compared with Gate 3 shows lower attention in the brain, and reduced extent of the low attention areas outside. Areas of high attention are concentrated towards the inferior and lateral background surrounding the brain.

### 4.7 GradCAM Activation Visualisation

Activation plots highlight areas of interest to the Critic. As shown in Figure 9, when scans are aligned to the template using affine registration (i.e. 12 degrees of freedom), in the generated images the Critic generally focuses on the white matter. In the target images (i.e. high-resolution images), the activation seems to be focused on the brain contour. The aqueduct also seems to be an area of attention in the target images (bottom left in Figure 9). When scans are aligned using rigid-body registration (i.e. 6 degrees of freedom), the pattern of activation is similar (Figure A.5).

**Figure 9:**
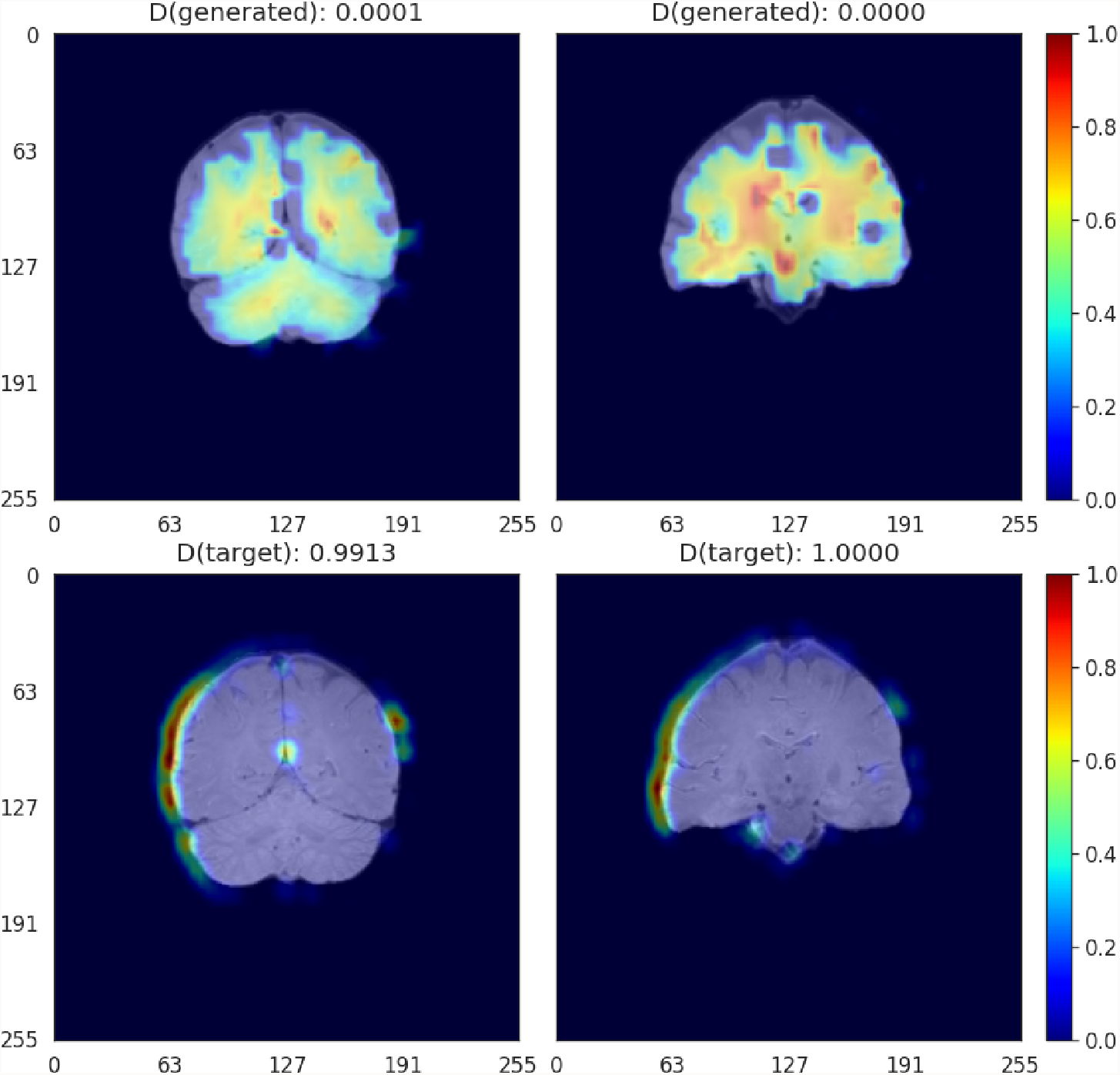
GradCAM activation plots for the Critic for affine-registered scans. Activation is represented in a `jet` colormap, with areas of high activation in red and low attention in blue.

### 4.8 Clinically Relevant Evaluation

#### 4.8.1 Brain Image Quality Metrics

Table 1 shows the results of subtracting the added quality metrics calculated from the upsampled images using Model 0 and Model 6, and the low resolution images resampled to the dimensions of the high resolution images using different types of interpolations and cost functions, from the quality metrics calculated from the target high resolution images. In general, with the exception of the EFC, in which the trilinear interpolation and the correlation ratio as cost function (i.e. default transformation used nowadays in clinical research) performed better for most of the protocols (i.e. 3 out of 4) represented in the testing subsample, in the rest of the metrics our SR reconstructed images were closed to the target high resolution images. Better GM/WM mean intensity ratio was obtained for the images from protocol 2, which comprised more than half of the testing set, upsampled using Model 0. Individual results from the different protocols for each of the three scan sequences that are included in our models can be seen in the Supplementary data spreadsheet.

**Table 1:** Mean Absolute Error (MAE) and standard deviation (STD) of the results of computing the anatomically meaningful metrics for upsampled and interpolated images against the high resolution images for each acquisition protocol in the testing subsample. Protocols 1 and 5 are represented in image data from only one patient (respectively). Two patient image data were acquired using protocol 3, and data from 5 patients are reflected in the results from protocol 2. N/A stands for Not Available, as STD can’t be calculated for data from only one or two sources. Best results are highlighted in bold.

| METRIC (PROTOCOL NO.) | MODEL 0 |  | MODEL 6 |  | INTERPOLATED SPLINE |  | INTERPOLATED SINC |  | INTERPOLATED TRILINEAR |  |
| --- | --- | --- | --- | --- | --- | --- | --- | --- | --- | --- |
|  | MAE | STD | MAE | STD | MAE | STD | MAE | STD | MAE | STD |
| EFC (1) | 18437 | N/A | **9480** | **N/A** | 53836 | N/A | 56046 | N/A | 28045 | N/A |
| EFC (2) | 76065 | 36119 | 99743 | 63486 | 65339 | 40743 | 65720 | 40366 | **58639** | **49944** |
| EFC (3) | 14188 | N/A | 12565 | N/A | 17625 | N/A | 17170 | N/A | **6318** | **N/A** |
| EFC (5) | 55260 | N/A | 45987 | N/A | 34107 | N/A | 33207 | N/A | **10418** | **N/A** |
| CJV (1) | 0.03499 | N/A | **0.02999** | **N/A** | 0.04869 | N/A | 0.04874 | N/A | 0.09952 | N/A |
| CJV (2) | **0.05463** | **0.05659** | 0.07352 | 0.03924 | 0.08816 | 0.04262 | 0.08856 | 0.04239 | 0.07706 | 0.05998 |
| CJV (3) | **0.02844** | **N/A** | 0.3429 | N/A | 0.1595 | N/A | 0.1610 | N/A | 0.08374 | N/A |
| CJV (5) | 0.01244 | N/A | **0.00580** | **N/A** | 0.01444 | N/A | 0.01443 | N/A | 0.1108 | N/A |
| GM/WM MEAN INTENSITY RATIO (1) | 0.02285 | N/A | 0.03899 | N/A | **0.01551** | **N/A** | 0.01641 | N/A | 0.08344 | N/A |
| GM/WM MEAN INTENSITY RATIO (2) | **0.03269** | **0.02544** | 0.07791 | 0.06963 | 0.09623 | 0.07512 | 0.09620 | 0.07480 | 0.09952 | 0.06232 |
| GM/WM MEAN INTENSITY RATIO (3) | 0.02627 | N/A | 0.0296 | N/A | 0.03597 | N/A | 0.03480 | N/A | **0.01541** | **N/A** |
| GM/WM MEAN INTENSITY RATIO (5) | 0.01237 | N/A | 0.00882 | N/A | 0.00939 | N/A | 0.00982 | N/A | **0.00600** | **N/A** |
| CSF/WM MEAN INTENSITY RATIO (1) | 0.1705 | N/A | **0.1469** | **N/A** | 0.3339 | N/A | 0.3319 | N/A | 0.2337 | N/A |
| CSF/WM MEAN INTENSITY RATIO (2) | **0.3133** | **0.3927** | 0.4501 | 0.5840 | 0.6675 | 0.8260 | 0.6665 | 0.8224 | 0.7128 | 0.8975 |
| CSF/WM MEAN INTENSITY RATIO (3) | 0.2110 | N/A | **0.2104** | **N/A** | 0.4060 | N/A | 0.4032 | N/A | 0.5501 | N/A |
| CSF/WM MEAN INTENSITY RATIO (5) | **0.1720** | **N/A** | 0.1790 | N/A | 0.3829 | N/A | 0.3819 | N/A | 0.4487 | N/A |

#### 4.8.2 Tissue Segmentation Accuracy - Results in the testing set

Volumes of the WMH, CSF, veins and dural meninges, total brain and pial layers of the reference high resolution images, the SR images from models 0 and 6, and the low resolution images resampled using different types of interpolation and cost functions, for each of the 5 test samples are given in the Supplementary Tables A.2-A.5. The same tables show the values of other similarity metrics that evaluate spatial agreement between the segmentation masks obtained in the high resolution sets against those in the upsampled. These are: Dice coefficient, true positive and negative fractions and positive predicted values.

Figure 10 shows the Bland-Altman plots of the agreement between three of the volumetric measurements obtained from the high-resolution images, and the images upsampled by different procedures, using the same scripts with identical (default) parameters. With the exception of two cases with protocols under-represented in the training set, for which the SR images obtained from our framework were of lower quality, the volumetric results from our segmentations differed from those obtained from the target images in a value ranging from 0 to 20-40% of the mean value between the two measurements. This is comparable (e.g. CSF segmentation) or better (e.g. venous sinuses and pial surfaces/brain-CSF boundaries/partial volume) than the measurements obtained from the upsampled images using any type of interpolation. Specifically the volume of the pial surfaces reported, as comprises CSF-brain parenchyma partial volume effects, is greatly affected by interpolated images as can be seen in the correspondent plot.

**Figure 10:**
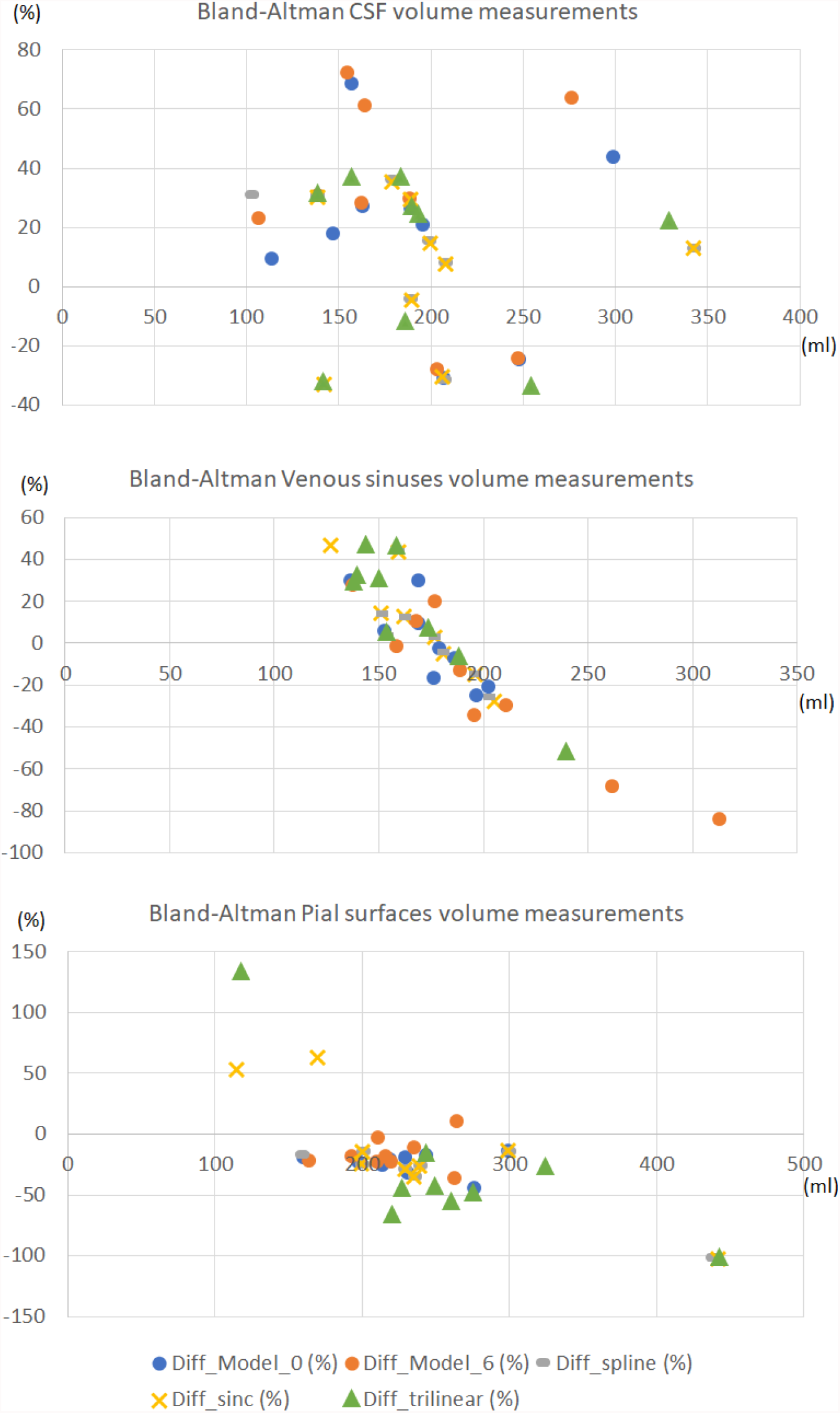
Bland-Altman plots of the agreement between the volumetric measurements obtained from the high-resolution images, and the images upsampled by different procedures, using the same scripts with identical parameters. The horizontal axes represents the average volume between the two volumetric measurements compared expressed in ml. The vertical axes represents the percentage difference between the volumetric measurement obtained from the upsampled image and the one obtained from the high-resolution (target) one, with respect to the mean value between the two measurements.

Figure 11 illustrates the segmentation masks of CSF-filled spaces, venous sinuses and dural meninges, and pial layers and membranes covering the brain parenchyma surface, superimposed in the FLAIR sequence, for one case, with a stroke lesion (i.e. hyperintensity) in the superior parietal cortex extending to the deep white matter visible in the coronal slice selected. The stroke is misclassified as pial structure in its entirety in the original high resolution image, and partially in the interpolated low resolution image, while correctly excluded from the masks shown in the reconstructed SR image. Normal tissues (grey (GM) and white matter (WM) masks not shown) yielded results comparable in accuracy between the high resolution and SR images, but run with different parameters and software. For example, while FSL-FAST run with default parameters yielded consistently good results segmenting GM and WM in high resolution images, this was not the case when the SR images were used as input, and priors needed to be given for it to perform well. Still in some cases, the SR T1-weighted images did not have enough contrast between GM and WM, and it was necessary to add anatomical knowledge to a multispectral segmentation (explained in Section 3.8.2 but considering more classes) developed in-house to obtain good results.

**Figure 11:**
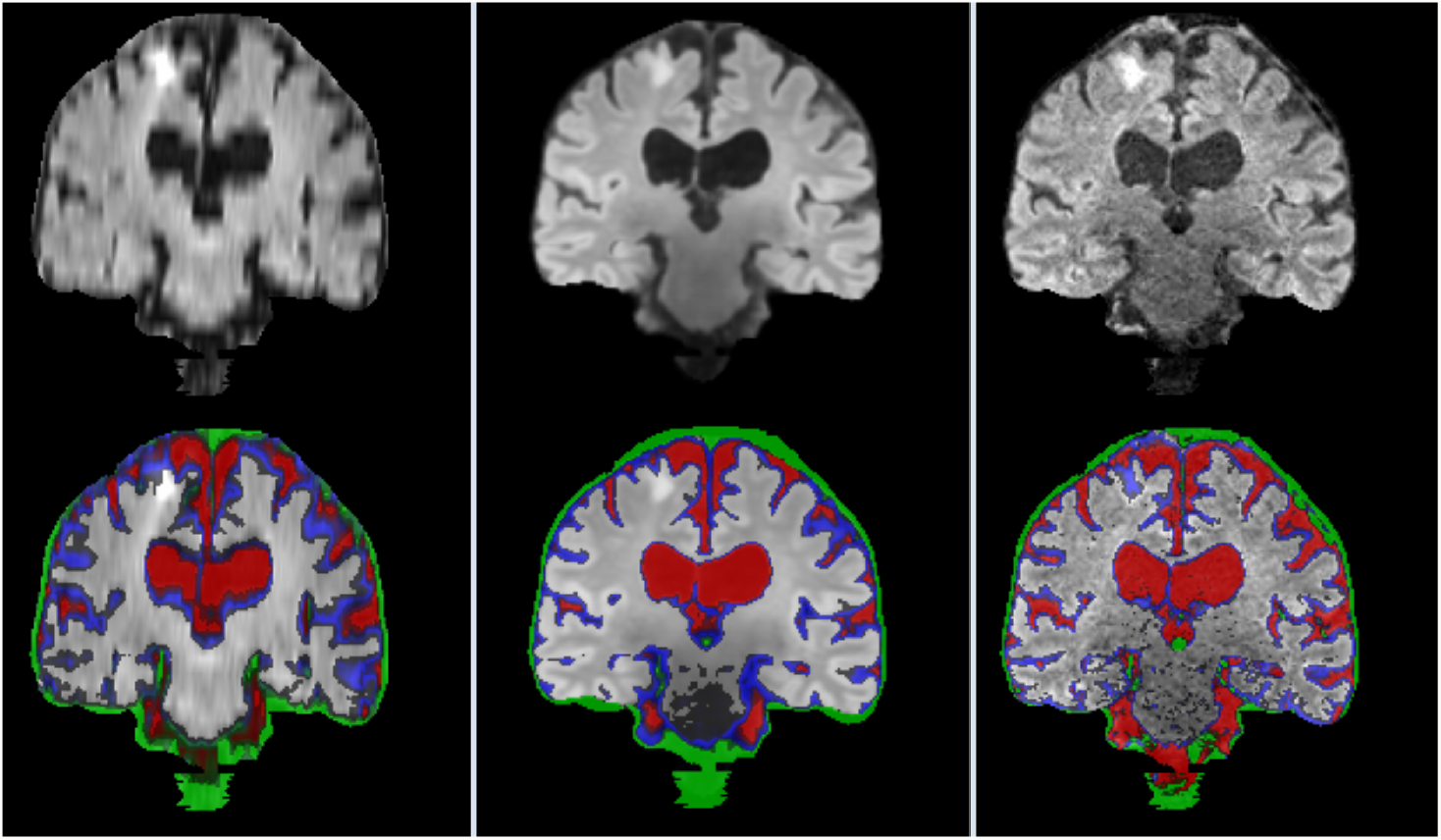
Coronal FLAIR slice of low resolution (left column) resampled and aligned to the high resolution image (right column) using trilinear interpolation, and the correspondent SR image (center column). Binary masks of CSF (red), pial layer (blue) and dural meninges and venous sinuses (green) are superimposed in the bottom row.

The results from manually editing the WMH segmentations obtained from using the high-resolution images spatially agreed better with the automatic results obtained from the upsampled scans than with those using interpolated images. Low Jaccard and Dice coefficients in general were in cases with prominent new and old stroke lesions manually removed from the reference segmentations, and partially included in the automated masks.

Figure 12 illustrates an axial FLAIR slice of a case in which the high-resolution image suffers from motion artefact and low SNR, and the low resolution FLAIR image has higher in-plane spatial resolution than the 3D-acquired images of the high resolution scan (voxel sizes of 0.47 × 0.47 × 6 mm3 vs. 0.973 × 0.973 × 0.973 mm3 respectively). As can be appreciated, the image upsampled with our SR scheme Model 0 has perceptually no motion artefacts and less noise compared to the high-resolution and less or none partial volume effect compared to the low-resolution; and the automatic WMH segmentation in the SR image has fewer false positives than the segmentation in both low- and high-resolution images. However, subtle lesions visible in the low resolution image are not reflected in the SR image, yielding few false negatives.

**Figure 12:**
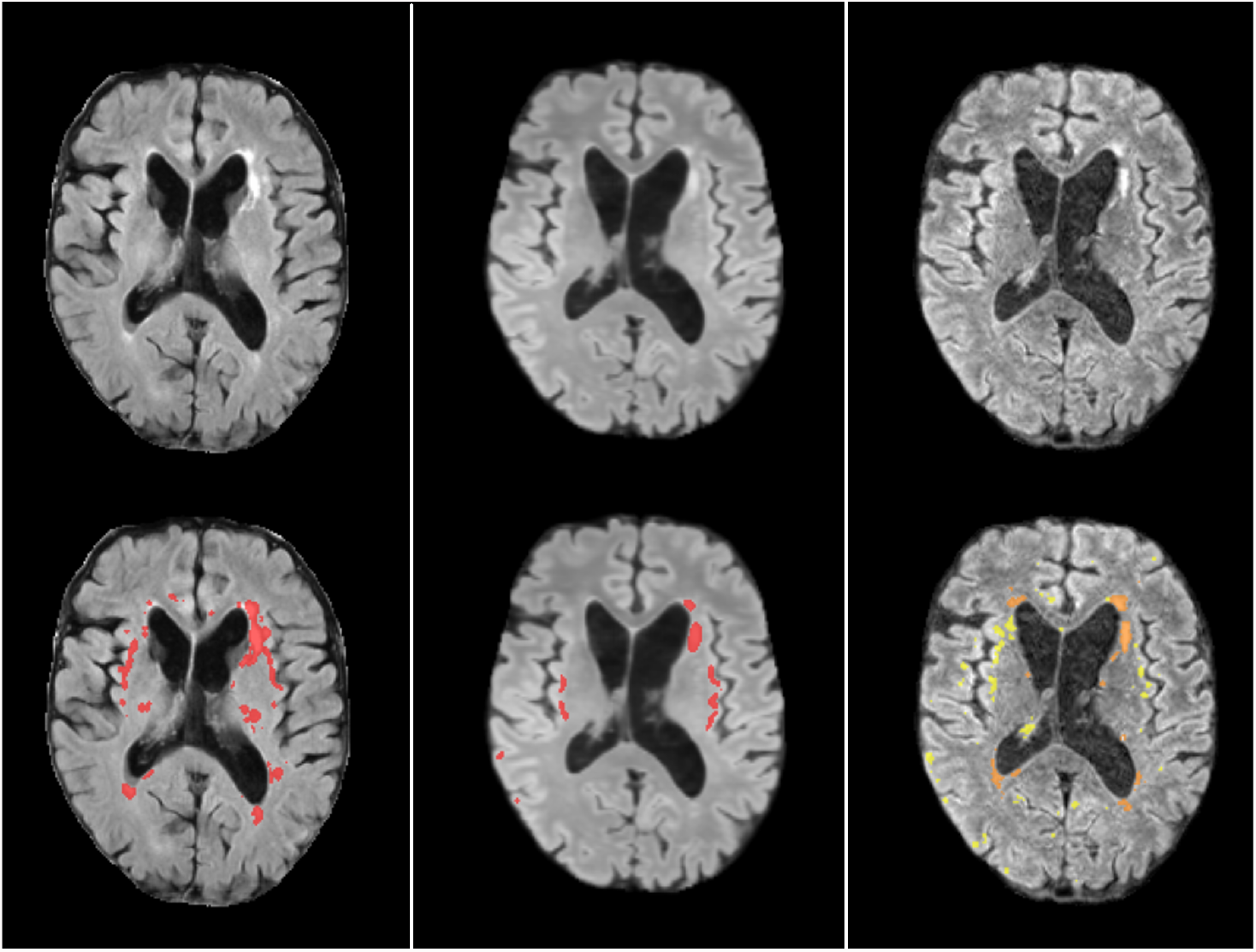
Axial FLAIR slice of low resolution (left column) resampled and aligned to the high resolution image (right column) using trilinear interpolation, and the correspondent SR image (center column). The bottom row shows the automatically generated binary masks of WMH superimposed in red for the low resolution and SR images, and in yellow in the high resolution image. The manually corrected WMH mask is also superimposed in the high resolution image and can be appreciated in orange tone.

#### 4.8.3 Tissue Segmentation Accuracy - Results in the control subsample

From the first 190 patients recruited in the primary study that provided data for the present analysis, 188 provided MRI data at baseline, 10 at 1 month, 24 at 2 months, 64 at 3 months, and 11 at 4 months. Longitudinal MRI data in this interval, all acquired with the same high resolution research protocol, was acquired for 87 patients. We analysed the variability in the segmentations on this subset to conclude on the plausibility of the differences found between the volumetric measurements of the upsampled images and their high-resolution target images acquired within a similar interval. Figure 13 shows the distribution of the volumetric measurements of the same tissue classes for which the agreement with the upsampled results is reflected in Figure 10, but not only for the baseline (reference) time point, but also at 1, 2, 3 and/or 4 months. Certainly fluctuations in the mean, median and quartiles at each month may be due to missing values, as not all participants were imaged at the same intervals, but subtraction of baseline volumes from those at subsequent time points showed median differences in the range of 1 to 4.72% [IQR from 0.91 to 8.21%] for CSF, 0.5 to 2.70% [IQR from 0.75 to 5.12%] for venous sinuses, and 9.24 to 12.71% [IQR from 7.60 to 14.27%] for pial surfaces/CSF-brain boundaries’ partial volume.

**Figure 13:**
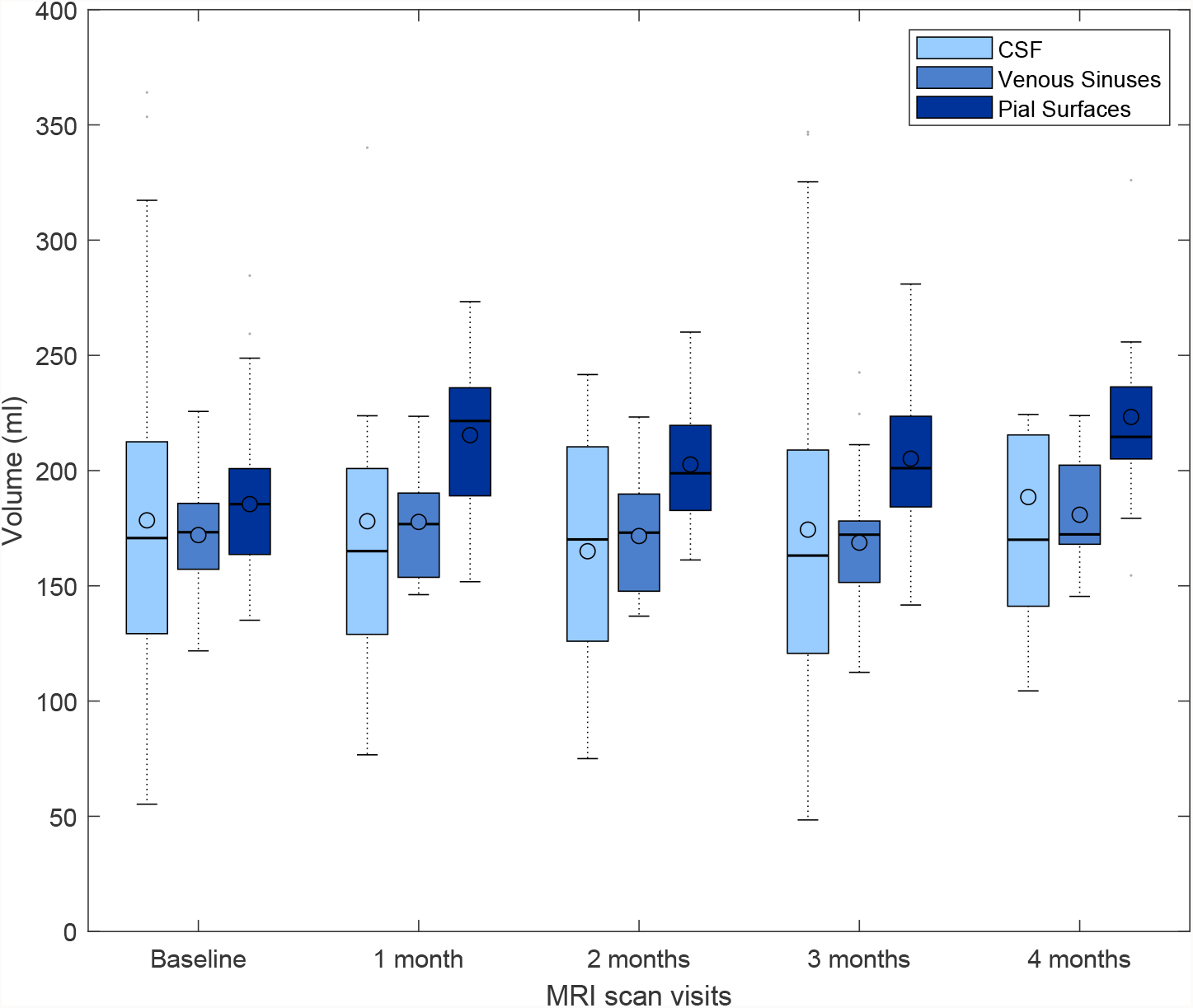
Box plots showing the full range, mean, median and interquartile range of the volumetric measurements of CSF-filled spaces, venous sinuses and pial layers/CFS-brain partial volume effects, for a subset of patients that underwent brain MRI scan at baseline, and subsequently, monthly up to 4 months after enrolling the primary study that provided data for the present analysis.

Figure A.6 shows the distribution (and variability) of the volumes of the venous sinuses (i.e. to be more stable across this small period of time) in the same subsample, across the same time intervals. As can be appreciated, the differences between baseline and subsequent measurements range up to approximately *±* 13%, similar to the variability in the agreement between measurements obtained from the upsampled and target images in the test sample.

#### 4.8.4 Visual Evaluation

Results of the visual evaluation can be appreciated in the supplementary file labelled as: `Li_Castorina_Valdes-Hernandez_et_al_Visual_evaluation.xlsx`. The scan manager established the following criteria for rating the images: 1) images pretty pixelated with no discernible disease markers, unsuitable for assessing them; 2) images quite fuzzy with barely visible disease markers, artefacts, most possibly unsuitable for assessing disease markers; 3) some fuzziness on images, visible disease markers such as lesions and lacunes, suitable for assessing them; 4) image quality satisfactory, very light blurriness, suitable for assessing disease markers; 5) high image quality, no blurriness, quite clear images, suitable for assessing disease markers.

As per the scan manager’s evaluation, the upsampled images for protocols 2 and 3 had superior or equal quality than the native images (i.e. native images have low spatial resolution at least in one plane), the results from protocol 5 were poor overall, and the dataset from protocol 1 saw a slight improvement only for the T1-weighted image, but the quality of the FLAIR and T2-weighted images from the patient data acquired with this protocol was considered being slightly downgraded in the upsampling.

The neurologist graded the images according to visibility of anatomical details and feasibility/clarity in the display of pathology. As per the neurologist’s evaluation, with the exception of the images from protocol 5, for which the upsampling produced results of poor quality, T1-weighted and FLAIR were considered of equal or improved quality after being upsampled, but all T2-weighted images ranked lower than the native images. In the two cases where the target high-resolution T2-weighted images were corrupted with artefacts, the upsampled T2-weighted were considered of superior quality with respect to them, although of equal or slightly inferior quality compared to the native images.

Overall, the majority of the SR images show all anatomical details of the high resolution images with high fidelity, with the exception of the images from the patient scanned with the protocol labelled as 5, which also was underrepresented in the training sample (i.e. 4/51 datasets). Target and native images corrupted by noise saw an upgrade on the results from using the framework proposed (average score 4/5).

## 5 Discussion

### 5.1 Upsampling Patches, Slices and Critic loss

Generally, upsampling whole slices rather than patches lead to better results. This may be due to the redundancy of the patches which may not cover the full image space whilst oversampling some specific areas.

Additionally, using a critic did not improve the model’s performance. We however believe that its utility goes beyond improving the accuracy, as it is a useful tool to identify potential artifacts in the upsampled images, thus increasing the explainability of the models. We propose the use of GradCAM to visualise the activations, highlighting potential artifacts, which may be useful as added information in clinical settings if the framework proposed is applied.

### 5.2 Upsampling MRI Sequences Separately and as Channels

Using data fusion of the three MRI structural sequences (i.e. FLAIR, T1-weighted, and T2-weighted), increased the performance of our model. As shown in Figure 5, information from one sequence is used in the upsampling/reconstruction of another sequence. In this multi-channel architecture, the convolution process considers more than just the one sequence/channel, thus allowing the flow of information between channels.

### 5.3 Interpreting Attention Gates

We proposed the use of attention gates to visualise the attention of the model at each step of the upsampling process as shown in Figure 8. We found a peculiar pattern, where the attention of Gates 2 and 4 was complementary to their respective previous gates, indicating that the network focuses on different areas of the images at different depths. It is likely that the network is learning to extract different levels of details at different gate dimensions. The lower gates might be differentiating the brain from the background while the higher gates might be focused on more specific details such as cortical folds and grey matter.

Additionally, Gate 2 and Gate 3 also cover areas of the brain which contain lesions related to SVD, thus further research could explore the effects of hyper-parameters and their effect on the quality of upsampled areas covering the disease-specific features.

We believe that attention gate plots are essential tools in the SR field to increase the explainability of the process and results. Currently, super-resolution models are understood as black-boxes, making them potentially sensitive to bias in the training data and, consequently reducing their usefulness in practical scenarios.

Further investigations of these maps may include channel-wise attention to differentiate attention for each sequence and understand in which extent one sequence, or an area of the sequence, is contributing towards the final upsampled image.

### 5.4 Insights from the Clinically Relevant Evaluation of the SR Framework’s Performance

#### 5.4.1 Datasets

The framework proposed yielded very good results for datasets acquired with protocols represented three or more times in the training set; especially in cases when the input low-resolution sequences of the scan were heterogeneously acquired, for example, one sequence acquired axially, other coronally and the other 3D or sagitally; or when the sequences were acquired in the same orientation but with different voxel sizes and spatial resolutions. When the images that conform the scans are homogeneously acquired in terms of orientation and spatial resolution, the framework proposed may have limited utility, given that the information missing in the low-resolution image sequences in their native space may not be synthesised properly.

Due to the nature of the clinical condition of the patients that provided imaging data (i.e. diagnostic scan acquired soon after presenting with mild stroke symptoms), and the resources required to obtained paired scans, the dataset of paired low- and high-resolution scans available was comprised from only 60 samples. This limits the amount of training data for our model and therefore its performance. Also, all scans available were from patients with sporadic SVD, who presented to a clinic with a mild to moderate stroke symptoms. Patients with other diseases/brain anomalies (e.g. tumours, large haemorrhages, brain injuries, Alzheimer’s or Huntington’s diseases, rare genetic neuropathies), or in a wider age range (e.g. children, adolescents) were not represented in the data. Therefore inaccuracies of a wider range than those discussed and presented in the Results would be expected if scans from other than sporadic SVD patients are input. With MRI scans becoming a more common practice, we expect more training data to be available in the future for re-training our model and improving its performance. Nevertheless, given that the most represented protocols in the training dataset are those used throughout the years in the two largest hospitals in the region, and the high incidence of sporadic SVD, this framework “as is” will allow the use of large amount of scan data for the development of robust AI-powered precision-medicine schemes.

#### 5.4.2 Representation of clinically relevant details

It is difficult to assert the extent in which the SR framework proposed reduces or enhances subtle clinically relevant details or not, given that the input and target images (used for training and testing) were acquired one month apart. As all data used was obtained from clinical settings, and resources were not available to allow both, high- and low-resolution scans, being acquired on the same day or in consecutive days, subtle lesional changes present in the low-resolution scan were sometimes absent in its SR reconstruction. As the low-resolution data was acquired soon after patients presented with stroke symptoms, subtle differences in the MRI are expected to occur in the interval elapsed between both scans. Results show that the SR framework proposed is useful in reconstructing scans that have been affected by image artefacts, which, otherwise have limited usefulness or even lack it. The proposed model could be applied, therefore, to reduce scan duration without compromising the quality of the scan when patients cannot tolerate longer scanning time.

#### 5.4.3 Feasibility for automated precision medicine applications

While the WMH, total brain tissue, and non-brain tissue structures and CSF-filled spaces could be consistently segmented with the same parameters using SR, original, and interpolated image data, allowing comparison of the results, this was not the case for normal tissues (i.e. WM and GM). These needed separate adjustments (i.e. priors and anatomical reference locations in a multispectral approach that used the three sequences) for yielding good results using SR images, thus contrasting with the good segmentations that are obtained from using only T1-weighted images acquired under research protocols as input to FSL-FAST, which is considered the brain tissue segmentation tool for excellence. Given the small size of the training dataset, and the low (or even absent) WM/GM contrast in some of the T1-weighted images acquired in clinical settings (i.e. used for training), this is not surprising. Re-training the model with more datasets could perhaps overcome this hurdle. However, in addition, it is worth to note that, for applying our CNN-based SR framework we linearly normalised the image intensities to double-precision values in the interval [0,1], so linearly rescaling back the intensities of the upsampled images to a full-scale 12-bit resolution integer values does not necessarily ensure fidelity in the intensity adjustment process. Future works can evaluate the gain (if any) in applying such more complex segmentation pipeline instead of using FSL-FAST with a single-sequence input, to the original and high-resolution scans.

### 5.5 Limitations and Future Work

In addition to the previously mentioned limitations related to representativeness and size of the data available, manual adjustment of the image contrast - a common process known as magnet shimming - visually guided, can also be a source of bias. Such process aims to reduce the influence of inhomogeneities in the static magnetic field (B0) of the scanner. But it might be inconsistent from person to person and inherently introduce a non-reproducible bias in the input data. In this work we also expect the low- and high-resolution scans to have the same dimension. This was done by either using a rigid, affine, or warping space transformations of the scans to a template. In future work, this pre-processing step can be optimised, and this optimisation be integrated as part of the data processing pipeline.

The fact that the template and high-resolution images had nearly 1 *mm*^3^ voxel dimensions, and some of the low-resolution images had double the in-plane resolution of the target images (despite having only few slices), influenced in the neurologist’s preference of the low-resolution scan for visual diagnostic assessment in some cases. Brain template and full scans with higher spatial resolution and isotropic voxels were not available. This limitation, however, reflects the present challenges that the whole fields of medical image analysis and precision medicine need to overcome in the near future. The use of transfer- or continuous-learning techniques with higher-resolution brain templates from higher-resolution isotropic brain scans will be beneficial not only to enhance the applicability of our framework, but to advance the whole field.

Another limitation is that the GradCAM activation map was averaged per channel, which is acceptable for identifying areas of interest to the model. However, for clinical use, using channel-wise localised attention map could trace the activation onto the specific MRI sequence that provides (versus lacks) the information the network is synthesising at each time and location. Similarly, this applies to our attention map obtained from the attention gates, meaning our model cannot learn fine-grained attention per sequence. Future work on channel-wise attention would identify specific areas of in the sequence relevant for the upsampled scan as well as further understanding the inter-channel information transfer. These will generally improve the explainability of the upsampled scans with clinically useful information. Future work could also explore the use of VGG perceptual loss as described by Johnson et al. [27] to enforce the performance to be reliant on the overall quality of the image rather than on pixel-wise metrics.

## 6 Conclusion

We have presented a framework for upsampling and reconstructing low resolution and low quality scans acquired in clinical practice. The proposed model shows promising for being applied to clinical research and precision medicine, and for adequately synthesising information from incomplete or corrupted sequences; making it a strong candidate for being applied in clinical settings to reduce scanning time without compromising the quality of the scan. Multi-channel data fusion and input data heterogeneity are essential to achieve quality in our framework’s output.

## Data Availability

All numerical data will be available upon publication.

## Acknowledgment

We would like to thank Justin Engelmann for his contribution in the early stages of this work. BML and LVC are supported by the United Kingdom Research and Innovation (grant EP/S02431X/1), UKRI Centre for Doctoral Training in Biomedical AI at the University of Edinburgh, School of Informatics. YC is supported by the China Scholarship Council. MCVH is funded by the Row Fogo Charitable Trust Grant no. BRO-D.FID3668413. SJW is funded by the Stroke Association Post-Doctoral Fellowship (SAPDF 18/ 100026). This work is also partially funded by the Selfridges Group Foundation under the Novel Biomarkers 2019 scheme (ref UB190097) administered by the Weston Brain Institute, and the Fondation Leducq Transatlantic Network of Excellence for the Study of Perivascular Spaces in Small Vessel Disease, ref no. 16 CVD 05. UC is funded by the Stroke Association Princess Margaret Research Development Fellowship 2018. FND is funded by the Stroke Association Garfield Weston Foundation Senior Clinical Lectureship(TSALECT 2015/04). DJG is Funded by the Wellcome Trust. The images used in this study were funded by the UK Dementia Research Institute which receives its funding from DRI Ltd, funded by the UK MRC, Alzheimer’s Society and Alzheimer’s Research UK. The 3T MRI Research scanner at The Royal Infirmary of Edinburgh, where the high-resolution images were acquired, is funded by the Wellcome Trust (104916/Z/14/Z), Dunhill Trust (R380R/1114), Edinburgh and Lothians Health Foundation (2012/17), Muir Maxwell Research Fund, Edinburgh Imaging, and The University of Edinburgh.

## A Appendix

### A.1 Detailed Model Configurations and Results

Table A.1 contains details of the settings used for each model evaluated as well as the performance metrics. Seed refers to the random seed used for better reproducibility, though GPU-accelerated model training can produce slightly different results depending on hardware.

**Table A.1:** Test performance for models in different configuration

| NAME | PRE-PROCESSING | $\lambda_{\text{CRITIC}}$ | LOSS | SR LOSS | MAE | NMSE | PSNR | SSIM |
| --- | --- | --- | --- | --- | --- | --- | --- | --- |
| 0 | AFFINE | 0 | BCE | 0.69592 | 0.003783 | **4.32E-10** | **35.39** | **0.9852** |
| 1 | AFFINE | 0 | BCE | 0.69600 | **0.003752** | 4.46E-10 | 35.25 | 0.9847 |
| 2 | AFFINE | 0 | BCE | 0.69601 | 0.003762 | 4.44E-10 | 35.29 | 0.9846 |
| 3 | AFFINE | 0 | MAE | 0.00372 | 0.003779 | 4.52E-10 | 35.21 | 0.9845 |
| 4 | AFFINE | 0 | BCE | 0.69596 | 0.003901 | 4.76E-10 | 34.92 | 0.9845 |
| 5 | AFFINE | 0 | BCE | 0.69601 | 0.003831 | 4.69E-10 | 35.04 | 0.9843 |
| 6 | RIGID | 0 | BCE | 0.69624 | 0.003878 | 4.35E-10 | 35.25 | 0.9842 |
| 7 | AFFINE | 0 | BCE | 0.69592 | 0.003796 | 4.65E-10 | 35.08 | 0.9841 |
| 8 | RIGID | 0 | BCE | 0.69626 | 0.003962 | 4.55E-10 | 35.07 | 0.9841 |
| 9 | RIGID | 0 | BCE | 0.69630 | 0.004009 | 4.68E-10 | 34.96 | 0.9839 |
| 10 | RIGID | 0 | BCE | 0.69627 | 0.004001 | 4.68E-10 | 34.98 | 0.9838 |
| 11 | RIGID | 0 | BCE | 0.69624 | 0.004044 | 4.68E-10 | 34.93 | 0.9837 |
| 12 | RIGID | 0.1 | BCE | 0.69621 | 0.004028 | 4.70E-10 | 34.93 | 0.9832 |
| 13 | AFFINE | 0 | BCE | 0.69606 | 0.004049 | 5.23E-10 | 34.59 | 0.9828 |
| 14 | RIGID | 0.5 | BCE | 0.69626 | 0.004109 | 4.72E-10 | 34.88 | 0.9824 |
| 15 | AFFINE | 0 | BCE | 0.69604 | 0.005163 | 6.63E-10 | 33.54 | 0.9770 |
| 16 | AFFINE | 0 | BCE | 0.69604 | 0.005337 | 7.05E-10 | 33.26 | 0.9758 |
| 17 | AFFINE | 0 | BCE | 0.69604 | 0.005656 | 7.18E-10 | 33.11 | 0.9735 |
| 18 | RIGID | 0.5 | BCE | 0.69547 | 0.012936 | 3.18E-09 | 26.54 | 0.9514 |
| 19 | AFFINE | 0.5 | BCE | 0.69666 | 0.014573 | 4.28E-09 | 25.33 | 0.9197 |
| 20 | WARP | 0 | MAE | 0.01962 | 0.021047 | 3.47E-09 | 26.52 | 0.8985 |
| 21 | WARP | 0 | BCE | 0.70339 | 0.020610 | 3.36E-09 | 26.64 | 0.8970 |
| 22 | WARP | 0.1 | BCE | 0.70327 | 0.022140 | 3.82E-09 | 26.07 | 0.8798 |

### A.2 Hyper-parameter Search Results

In Figure A.1, we tested the effect of varying patch size: 8 × 8, 16 × 16, 32 × 32, 48 × 48, and 64 × 64 on the performance metrics `PSNR`, `SSIM` and `MAE`. We also present the same patch dimensions using each MRI sequence as a channel (merge). Generally, higher patch dimensions tend to give better results when each sequence is considered separately, while when merged, the performance is better and the patch dimension becomes less important.

In Figure A.2 and A.3, we test the effect of varying number of patches and patch dimension on performance metrics, merging each sequence as a channel (merge) and separately (split) respectively. Generally the higher the number of patches, the higher thee performance at every patch dimension.

Finally, in Figure A.4 we test the effect of varying filter size on performance metrics for both merge and split sequence channels, and using the full scan or patches. Generally, higher number of filters improves the performance slightly, after about 64. Merge and full scan is the best performing combination in terms of `PSNR`, `SSIM` and `MAE`.

### A.3 Activation Visualisation

In Figure A.5, we show the GradCAM of the critic for the best performing models, Model 0 and Model 6 for Affine and Rigid pre-processing.

### A.4 Image Segmentation Results

Tables A.2, A.3, A.4, and A.5 show the volumetric results of the segmentation and the similarity metrics between the results from using each of the upsampled images against those obtained from segmenting the high resolution scan.

**Figure A.1:**
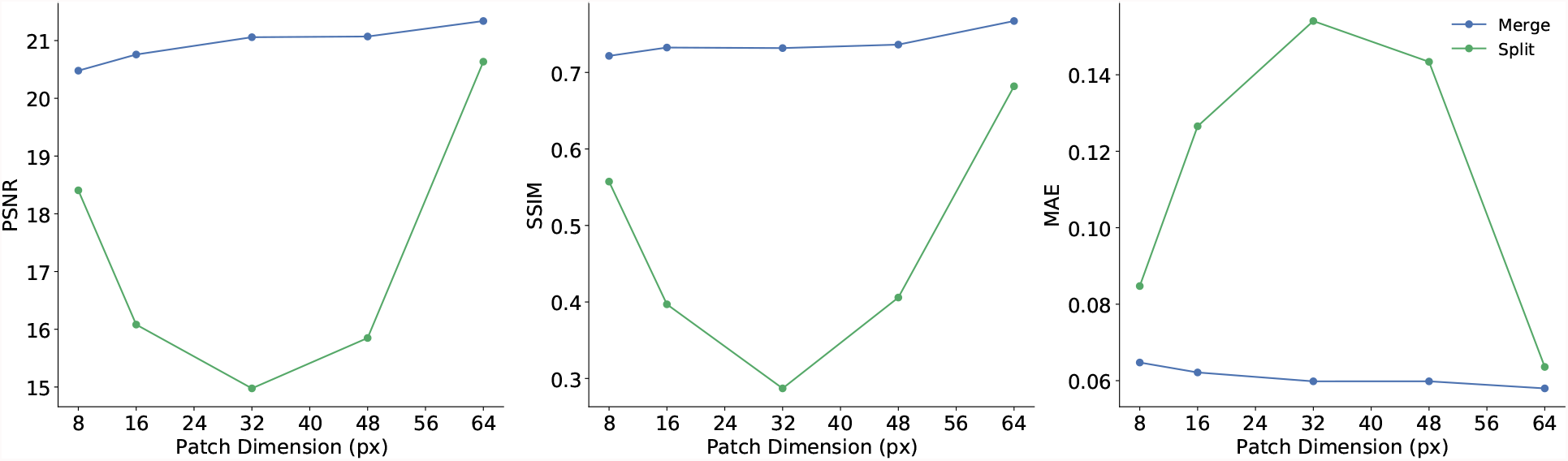
Performance metrics for merged (i.e. merged T1-weighted, T2-weighted, FLAIR channels) and split (T1-weighted, T2-weighted, FLAIR input separately) with increasingly larger patch dimensions

**Figure A.2:**
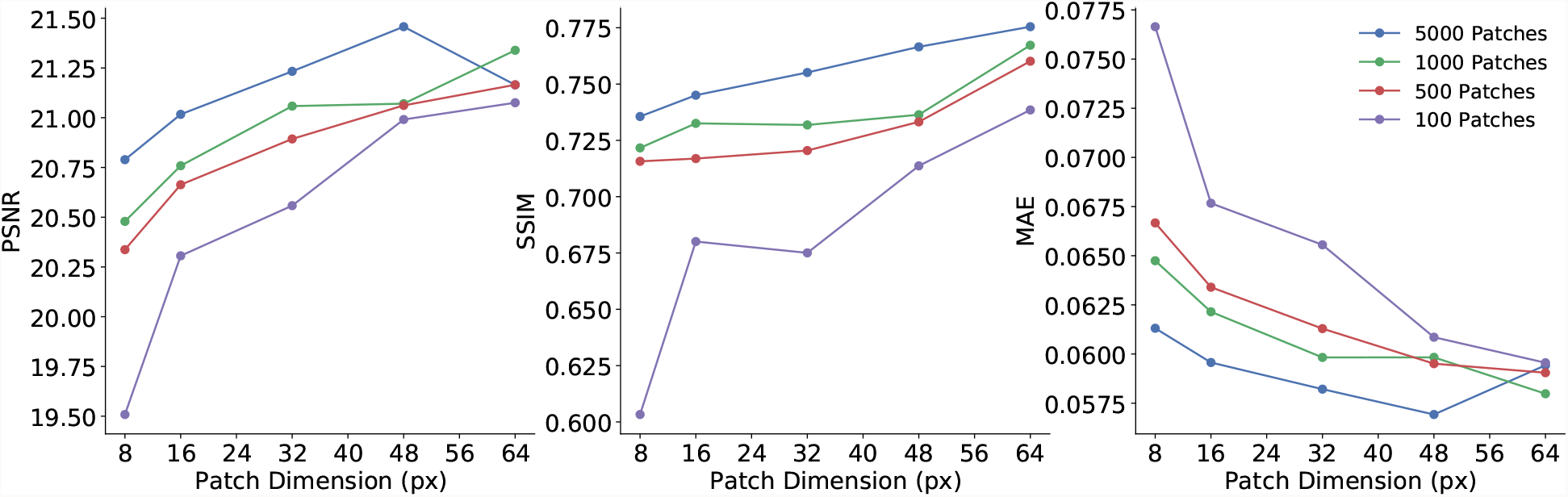
Performance metrics for merged scan sequences (i.e. data fusion of T1-weighted, T2-weighted, FLAIR channels) using different number of patches.

“Upsampled affine” refers to the SR scan sequences obtained from Model 0, “upsampled rigid” refers to the SR scan sequences obtained from Model 6, “interpolated spline” refers to the scan upsampled using spline interpolation and mutual information as cost function, “interpolated sinc” refers to the scan upsampled using sinc interpolation and mutual information as cost function, and “interpolated trilinear” refers to the scan upsampled using trilinear interpolation and the correlation ratio as cost function (default).

**Figure A.3:**
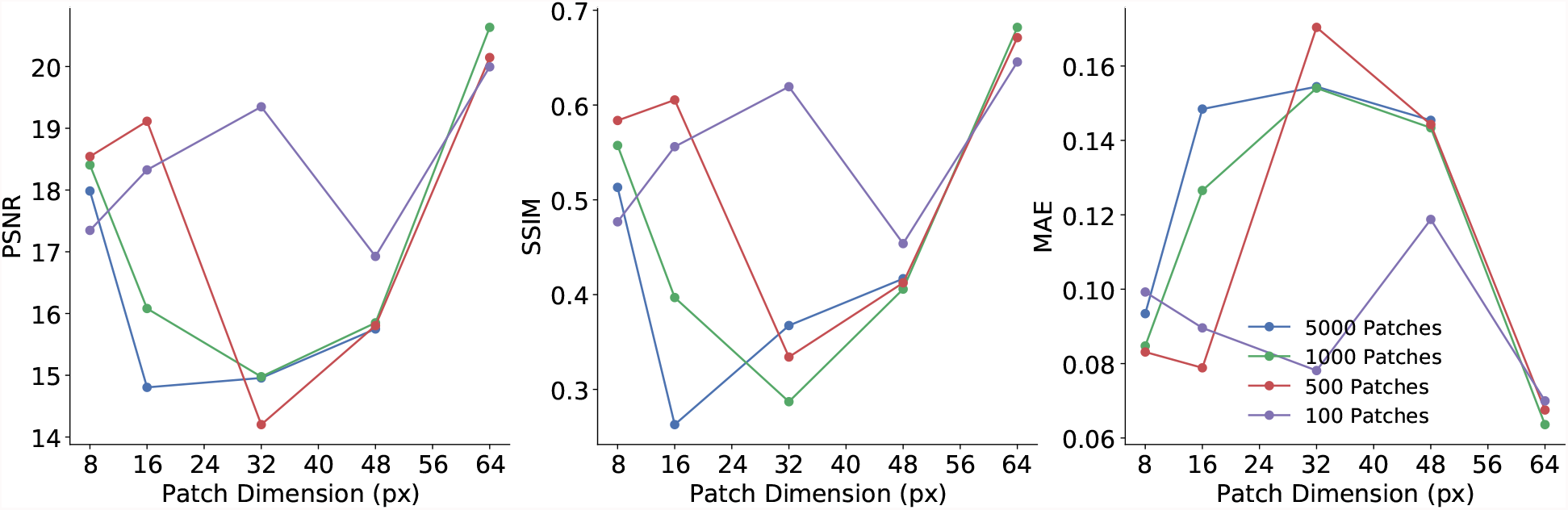
Performance metrics for scan sequences input separately (i.e. T1-weighted, T2-weighted, FLAIR treated independently) using different number of patches.

**Figure A.4:**
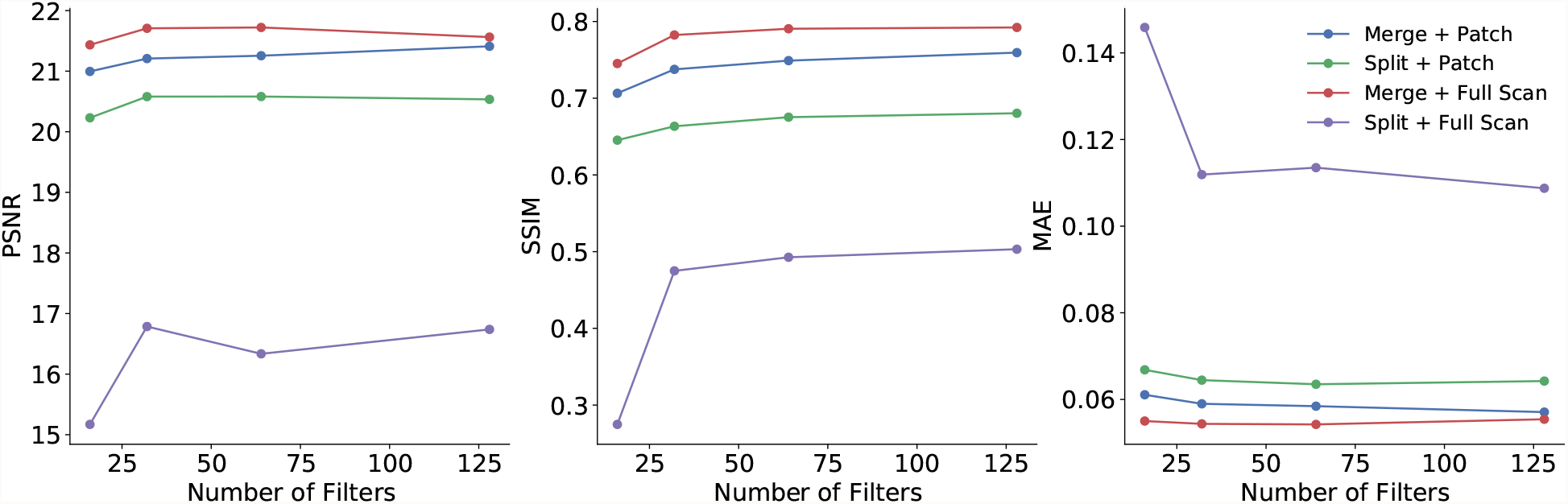
Performance metrics for different combinations of fused (T1-weighted, T2-weighted, and FLAIR channels) and split (T1-weighted, T2-weighted, and FLAIR treated separately) input image data entered either as full slices or in patches, in experiments that used different number of filters.

**Figure A.5:**
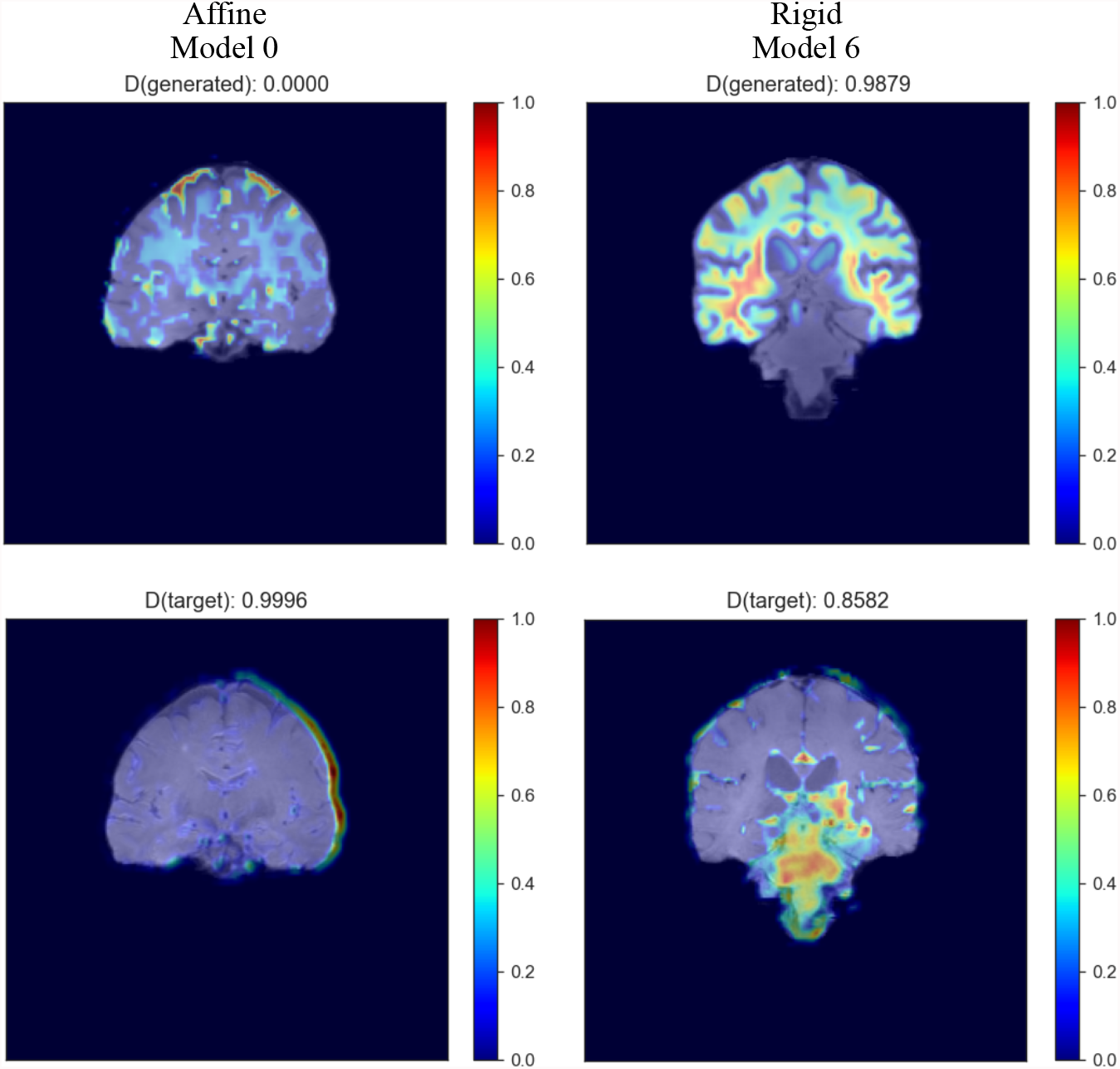
Example of GradCAM activation plots of the critic in (Left) Model 0 and (Right) Model 6, for images co-registered using Affine and Rigid-body registration respectively. Activation is represented in a `jet` colormap, with areas of high activation in red and low attention in blue.

**Figure A.6:**
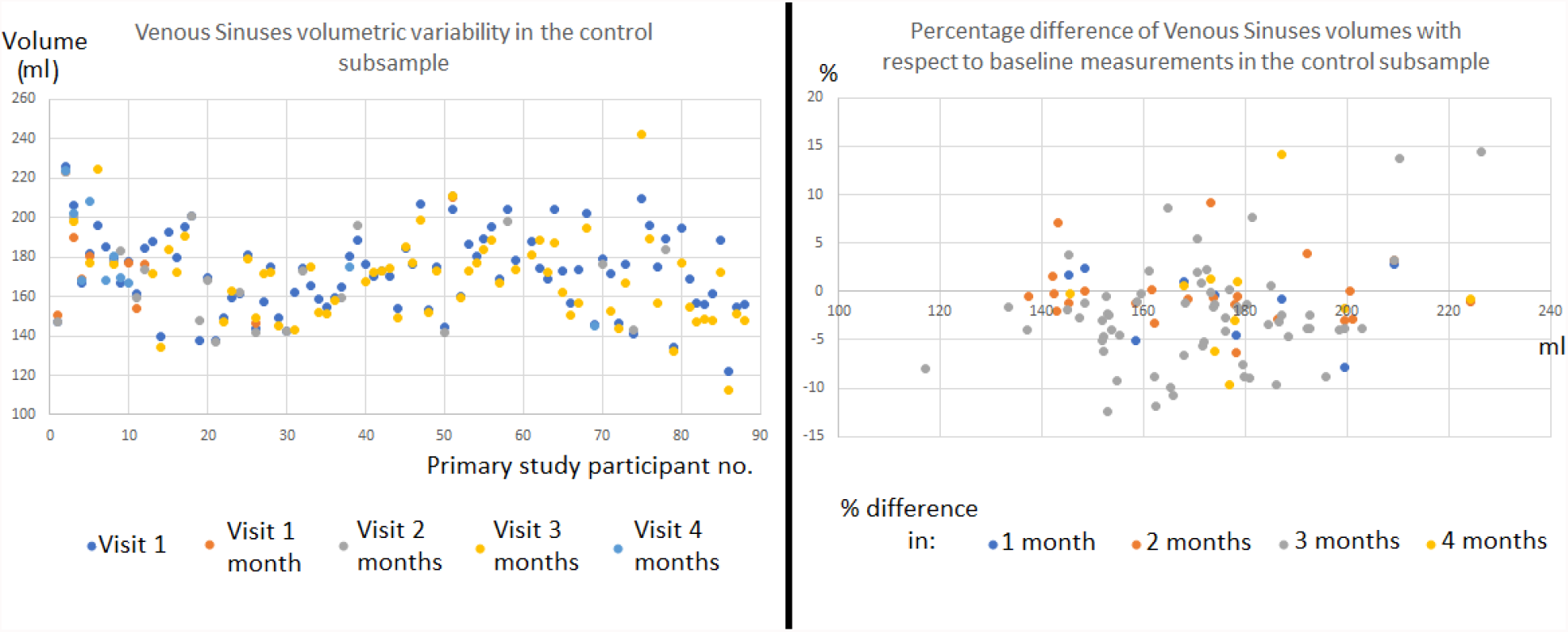
Volumetric variability of the venous sinuses in the control subsample throughout the 4 months after enrolling in the primary study that provided data for the present analysis.

**Table A.2:** White matter hyperintensties (WMH) volume measurements and metrics in the testing set (five patients’ data) reflecting the spatial agreement between WMH binary masks segmented in upsampled and high resolution scans. Dc: Dice coefficient, TPF: true positive fraction, PPV: positive predicted value, and TNF: true negative fraction.

| GROUP | PATIENT_NO | REF_VOL_ML | WMH_VOL_ML | WMH_Dc | WMH_TPF | WMH_PPV | WMH_TNF |
| --- | --- | --- | --- | --- | --- | --- | --- |
| UPSAMPLED AFFINE | 2 | 12.856 | 5.036 | 0.171 | 0.119 | 0.304 | 0.998 |
|  | 9 | 3.231 | 7.939 | 0.216 | 0.373 | 0.152 | 0.996 |
|  | 14 | 5.283 | 3.692 | 0.502 | 0.427 | 0.611 | 0.999 |
|  | 35 | 7.451 | 4.311 | 0.418 | 0.330 | 0.570 | 0.999 |
|  | 38 | 37.444 | 28.230 | 0.763 | 0.669 | 0.887 | 0.998 |
|  | 58 | 1.899 | 5.757 | 0.045 | 0.091 | 0.030 | 0.997 |
|  | 59 | 29.540 | 28.213 | 0.540 | 0.527 | 0.552 | 0.992 |
|  | 60 | 2.218 | 10.674 | 0.113 | 0.327 | 0.068 | 0.995 |
|  | 61 | 7.475 | 17.896 | 0.251 | 0.425 | 0.178 | 0.991 |
| UPSAMPLED RIGID | 2 | 12.856 | 4.028 | 0.195 | 0.128 | 0.408 | 0.999 |
|  | 9 | 3.231 | 5.309 | 0.178 | 0.235 | 0.143 | 0.997 |
|  | 14 | 5.283 | 4.004 | 0.471 | 0.414 | 0.547 | 0.999 |
|  | 35 | 7.451 | 2.925 | 0.365 | 0.254 | 0.647 | 0.999 |
|  | 38 | 37.444 | 29.020 | 0.768 | 0.682 | 0.880 | 0.998 |
|  | 58 | 1.899 | 7.383 | 0.051 | 0.124 | 0.032 | 0.996 |
|  | 59 | 29.540 | 16.088 | 0.535 | 0.413 | 0.758 | 0.998 |
|  | 60 | 2.218 | 7.799 | 0.064 | 0.144 | 0.041 | 0.996 |
|  | 61 | 7.475 | 14.558 | 0.263 | 0.387 | 0.199 | 0.993 |
| INTERPOLATED SPLINE | 2 | 12.856 | 30.130 | 0.232 | 0.388 | 0.166 | 0.986 |
|  | 9 | 3.231 | 20.116 | 0.153 | 0.552 | 0.089 | 0.989 |
|  | 14 | 5.283 | 13.294 | 0.365 | 0.642 | 0.255 | 0.994 |
|  | 35 | 7.451 | 16.166 | 0.319 | 0.505 | 0.233 | 0.992 |
|  | 38 | 37.444 | 56.467 | 0.643 | 0.806 | 0.535 | 0.983 |
|  | 58 | 1.899 | 3.787 | 0.112 | 0.167 | 0.084 | 0.998 |
|  | 59 | 29.540 | 35.995 | 0.619 | 0.687 | 0.563 | 0.990 |
|  | 60 | 2.218 | 6.005 | 0.095 | 0.176 | 0.065 | 0.997 |
|  | 61 | 7.475 | - | - | - | - | - |
| INTERPOLATED SINC | 2 | 12.856 | 29.758 | 0.233 | 0.387 | 0.167 | 0.987 |
|  | 9 | 3.231 | 19.872 | 0.154 | 0.550 | 0.089 | 0.989 |
|  | 14 | 5.283 | 13.147 | 0.366 | 0.639 | 0.257 | 0.995 |
|  | 35 | 7.451 | 16.012 | 0.318 | 0.501 | 0.233 | 0.993 |
|  | 38 | 37.444 | 56.087 | 0.645 | 0.805 | 0.538 | 0.983 |
|  | 58 | 1.899 | 3.830 | 0.111 | 0.167 | 0.083 | 0.998 |
|  | 59 | 29.540 | 35.668 | 0.620 | 0.685 | 0.567 | 0.990 |
|  | 60 | 2.218 | 5.975 | 0.095 | 0.176 | 0.065 | 0.997 |
|  | 61 | 7.475 | - | - | - | - | - |
| INTERPOLATED TRILINEAR | 2 | 12.856 | 39.926 | 0.213 | 0.437 | 0.141 | 0.982 |
|  | 9 | 3.231 | 28.574 | 0.123 | 0.606 | 0.069 | 0.983 |
|  | 14 | 5.283 | 25.339 | 0.257 | 0.745 | 0.155 | 0.988 |
|  | 35 | 7.451 | 22.941 | 0.268 | 0.546 | 0.178 | 0.988 |
|  | 38 | 37.444 | 69.461 | 0.585 | 0.835 | 0.450 | 0.975 |
|  | 58 | 1.899 | 3.613 | 0.117 | 0.170 | 0.089 | 0.998 |
|  | 59 | 29.540 | 38.948 | 0.608 | 0.705 | 0.534 | 0.988 |
|  | 60 | 2.218 | 7.924 | 0.086 | 0.197 | 0.055 | 0.996 |
|  | 61 | 7.475 | 16.480 | 0.188 | 0.302 | 0.137 | 0.991 |

**Table A.3:** Cerebrospinal fluid (CSF) volume measurements and metrics in the testing set (five patients’ data) reflecting the spatial agreement between CSF binary masks segmented in upsampled and high resolution scans. Dc: Dice coefficient, TPF: true positive fraction, PPV: positive predicted value, and TNF: true negative fraction.

| GROUP | PATIENT_NO | REF_VOL_ML | CSF_VOL_ML | CSF_Dc | CSF_TPF | CSF_PPV | CSF_TNF |
| --- | --- | --- | --- | --- | --- | --- | --- |
| UPSAMPLED AFFINE | 2 | 364.132 | 232.727 | 0.744 | 0.610 | 0.954 | 0.993 |
|  | 9 | 215.991 | 175.225 | 0.799 | 0.724 | 0.892 | 0.986 |
|  | 14 | 214.384 | 164.068 | 0.766 | 0.677 | 0.884 | 0.988 |
|  | 35 | 159.727 | 133.431 | 0.742 | 0.681 | 0.815 | 0.983 |
|  | 38 | 185.265 | 140.967 | 0.759 | 0.668 | 0.878 | 0.988 |
|  | 58 | 118.644 | 108.066 | 0.642 | 0.613 | 0.673 | 0.976 |
|  | 59 | 210.920 | 103.028 | 0.502 | 0.374 | 0.765 | 0.983 |
|  | 60 | 174.790 | 238.210 | 0.738 | 0.872 | 0.640 | 0.951 |
|  | 61 | 217.245 | 277.701 | 0.698 | 0.795 | 0.622 | 0.922 |
| UPSAMPLED RIGID | 2 | 364.132 | 187.627 | 0.652 | 0.494 | 0.959 | 0.995 |
|  | 9 | 215.991 | 160.157 | 0.761 | 0.662 | 0.893 | 0.988 |
|  | 14 | 214.384 | 113.870 | 0.622 | 0.476 | 0.897 | 0.993 |
|  | 35 | 159.727 | 116.733 | 0.702 | 0.608 | 0.831 | 0.987 |
|  | 38 | 185.265 | 139.044 | 0.752 | 0.658 | 0.877 | 0.988 |
|  | 58 | 118.644 | 94.025 | 0.612 | 0.549 | 0.692 | 0.981 |
|  | 59 | 210.920 | 98.705 | 0.485 | 0.356 | 0.761 | 0.983 |
|  | 60 | 174.790 | 231.317 | 0.740 | 0.860 | 0.651 | 0.954 |
|  | 61 | 217.245 | 277.178 | 0.695 | 0.791 | 0.620 | 0.922 |
| INTERPOLATED SPLINE | 2 | 364.132 | 320.600 | 0.672 | 0.631 | 0.717 | 0.940 |
|  | 9 | 215.991 | 199.312 | 0.679 | 0.652 | 0.707 | 0.958 |
|  | 14 | 214.384 | 183.446 | 0.576 | 0.535 | 0.623 | 0.956 |
|  | 35 | 159.727 | 117.281 | 0.540 | 0.468 | 0.638 | 0.971 |
|  | 38 | 185.265 | 193.113 | 0.564 | 0.576 | 0.552 | 0.938 |
|  | 58 | 118.644 | 86.912 | 0.545 | 0.472 | 0.645 | 0.979 |
|  | 59 | 210.920 | 146.631 | 0.595 | 0.504 | 0.726 | 0.971 |
|  | 60 | 174.790 | 240.398 | 0.684 | 0.813 | 0.591 | 0.944 |
|  | 61 | 217.245 | - | - | - | - | - |
| INTERPOLATED SINC | 2 | 364.132 | 319.407 | 0.670 | 0.629 | 0.717 | 0.940 |
|  | 9 | 215.991 | 199.713 | 0.677 | 0.652 | 0.705 | 0.957 |
|  | 14 | 214.384 | 184.945 | 0.576 | 0.536 | 0.622 | 0.955 |
|  | 35 | 159.727 | 117.698 | 0.540 | 0.469 | 0.637 | 0.971 |
|  | 38 | 185.265 | 193.544 | 0.564 | 0.576 | 0.552 | 0.938 |
|  | 58 | 118.644 | 165.748 | 0.569 | 0.682 | 0.488 | 0.943 |
|  | 59 | 210.920 | 147.327 | 0.595 | 0.505 | 0.724 | 0.971 |
|  | 60 | 174.790 | 237.450 | 0.684 | 0.807 | 0.594 | 0.945 |
|  | 61 | 217.245 | 161.164 | 0.495 | 0.431 | 0.581 | 0.950 |
| INTERPOLATED TRILINEAR | 2 | 364.132 | 293.370 | 0.692 | 0.625 | 0.776 | 0.956 |
|  | 9 | 215.991 | 169.770 | 0.698 | 0.623 | 0.793 | 0.975 |
|  | 14 | 214.384 | 164.250 | 0.612 | 0.540 | 0.705 | 0.969 |
|  | 35 | 159.727 | 117.064 | 0.564 | 0.488 | 0.666 | 0.974 |
|  | 38 | 185.265 | 128.168 | 0.602 | 0.509 | 0.736 | 0.976 |
|  | 58 | 118.644 | 164.861 | 0.587 | 0.701 | 0.505 | 0.945 |
|  | 59 | 210.920 | 297.714 | 0.634 | 0.765 | 0.542 | 0.901 |
|  | 60 | 174.790 | 197.704 | 0.691 | 0.737 | 0.651 | 0.961 |
|  | 61 | 217.245 | 150.216 | 0.580 | 0.491 | 0.710 | 0.968 |

**Table A.4:** Venous sinuses and meningeal dura volume measurements and metrics in the testing set (five patients’ data) reflecting the spatial agreement between the binary masks segmented in upsampled and high resolution scans. Dc: Dice coefficient, TPF: true positive fraction, PPV: positive predicted value, and TNF: true negative fraction.

| GROUP | PATIENT_NO | REF_VOL_ML | MEN_VOL_ML | MEN_DC | MEN_TPF | EN_PPV | MEN_TNF |
| --- | --- | --- | --- | --- | --- | --- | --- |
| UPSAMPLED AFFINE | 2 | 181.487 | 223.698 | 0.788 | 0.879 | 0.713 | 0.962 |
|  | 9 | 161.764 | 190.394 | 0.783 | 0.852 | 0.724 | 0.963 |
|  | 14 | 172.399 | 220.934 | 0.775 | 0.884 | 0.690 | 0.957 |
|  | 35 | 179.581 | 192.782 | 0.796 | 0.825 | 0.769 | 0.969 |
|  | 38 | 176.610 | 180.845 | 0.799 | 0.808 | 0.789 | 0.973 |
|  | 58 | 156.837 | 116.055 | 0.735 | 0.640 | 0.864 | 0.989 |
|  | 59 | 194.377 | 143.561 | 0.635 | 0.552 | 0.747 | 0.974 |
|  | 60 | 176.736 | 160.870 | 0.798 | 0.762 | 0.837 | 0.985 |
|  | 61 | 157.301 | 148.411 | 0.742 | 0.721 | 0.764 | 0.975 |
| UPSAMPLED RIGID | 2 | 181.487 | 443.607 | 0.529 | 0.911 | 0.323 | 0.835 |
|  | 9 | 161.764 | 228.804 | 0.706 | 0.852 | 0.602 | 0.937 |
|  | 14 | 172.399 | 350.651 | 0.601 | 0.911 | 0.448 | 0.880 |
|  | 35 | 179.581 | 242.252 | 0.705 | 0.829 | 0.614 | 0.936 |
|  | 38 | 176.610 | 201.023 | 0.761 | 0.814 | 0.715 | 0.959 |
|  | 58 | 156.837 | 118.099 | 0.741 | 0.649 | 0.862 | 0.989 |
|  | 59 | 194.377 | 158.523 | 0.650 | 0.590 | 0.723 | 0.969 |
|  | 60 | 176.736 | 158.898 | 0.809 | 0.768 | 0.855 | 0.987 |
|  | 61 | 157.301 | 159.777 | 0.757 | 0.763 | 0.751 | 0.972 |
| INTERPOLATED SPLINE | 2 | 181.487 | 210.247 | 0.705 | 0.761 | 0.657 | 0.957 |
|  | 9 | 161.764 | 140.938 | 0.718 | 0.672 | 0.771 | 0.978 |
|  | 14 | 172.399 | 152.347 | 0.739 | 0.696 | 0.787 | 0.980 |
|  | 35 | 179.581 | 174.318 | 0.747 | 0.736 | 0.758 | 0.971 |
|  | 38 | 176.610 | 184.908 | 0.668 | 0.683 | 0.653 | 0.954 |
|  | 58 | 156.837 | 151.997 | 0.769 | 0.757 | 0.782 | 0.977 |
|  | 59 | 194.377 | 124.921 | 0.681 | 0.560 | 0.871 | 0.989 |
|  | 60 | 176.736 | 229.035 | 0.659 | 0.757 | 0.584 | 0.946 |
|  | 61 | 157.301 | - | - | - | - | - |
| INTERPOLATED SINC | 2 | 181.487 | 210.272 | 0.704 | 0.760 | 0.656 | 0.957 |
|  | 9 | 161.764 | 140.284 | 0.716 | 0.669 | 0.771 | 0.978 |
|  | 14 | 172.399 | 151.682 | 0.737 | 0.693 | 0.788 | 0.980 |
|  | 35 | 179.581 | 174.328 | 0.746 | 0.735 | 0.757 | 0.971 |
|  | 38 | 176.610 | 185.344 | 0.666 | 0.682 | 0.650 | 0.954 |
|  | 58 | 156.837 | 97.213 | 0.646 | 0.523 | 0.844 | 0.990 |
|  | 59 | 194.377 | 124.537 | 0.680 | 0.558 | 0.871 | 0.989 |
|  | 60 | 176.736 | 233.256 | 0.655 | 0.759 | 0.575 | 0.944 |
|  | 61 | 157.301 | - | - | - | - | - |
| INTERPOLATED TRILINEAR | 2 | 181.487 | 194.894 | 0.611 | 0.634 | 0.590 | 0.953 |
|  | 9 | 161.764 | 117.529 | 0.633 | 0.546 | 0.752 | 0.980 |
|  | 14 | 219.860 | 127.085 | 0.621 | 0.540 | 0.732 | 0.979 |
|  | 35 | 179.581 | 167.828 | 0.722 | 0.698 | 0.747 | 0.971 |
|  | 38 | 176.610 | 110.496 | 0.594 | 0.483 | 0.772 | 0.982 |
|  | 58 | 156.837 | 150.297 | 0.771 | 0.755 | 0.788 | 0.978 |
|  | 59 | 194.377 | 122.331 | 0.674 | 0.549 | 0.873 | 0.989 |
|  | 60 | 176.736 | 301.850 | 0.560 | 0.758 | 0.444 | 0.905 |
|  | 61 | 157.301 | 118.263 | 0.650 | 0.569 | 0.757 | 0.980 |

**Table A.5:** Pial volume measurements and metrics in the testing set (five patients’ data) reflecting the spatial agreement between the binary masks segmented in upsampled and high resolution scans. Dc: Dice coefficient, TPF: true positive fraction, PPV: positive predicted value, and TNF: true negative fraction.

| GROUP | PATIENT_NO | REF_VOL_ML | MEN_VOL_ML | MEN_DC | MEN_TPF | MEN_PPV | MEN_TNF |
| --- | --- | --- | --- | --- | --- | --- | --- |
| UPSAMPLED AFFINE | 2 | 278.666 | 319.910 | 0.547 | 0.587 | 0.512 | 0.902 |
|  | 9 | 175.101 | 219.451 | 0.580 | 0.654 | 0.522 | 0.926 |
|  | 14 | 207.782 | 250.511 | 0.613 | 0.676 | 0.560 | 0.930 |
|  | 35 | 194.529 | 266.621 | 0.583 | 0.691 | 0.504 | 0.908 |
|  | 38 | 186.268 | 240.172 | 0.596 | 0.682 | 0.529 | 0.919 |
|  | 58 | 145.504 | 175.274 | 0.428 | 0.472 | 0.392 | 0.927 |
|  | 59 | 196.177 | 241.084 | 0.320 | 0.357 | 0.290 | 0.878 |
|  | 60 | 215.893 | 336.770 | 0.429 | 0.548 | 0.352 | 0.873 |
|  | 61 | 222.737 | 263.702 | 0.375 | 0.409 | 0.346 | 0.871 |
| UPSAMPLED RIGID | 2 | 278.666 | 249.163 | 0.516 | 0.488 | 0.546 | 0.929 |
|  | 9 | 175.101 | 210.189 | 0.570 | 0.627 | 0.522 | 0.930 |
|  | 14 | 207.782 | 213.674 | 0.570 | 0.578 | 0.562 | 0.941 |
|  | 35 | 194.529 | 243.904 | 0.567 | 0.639 | 0.509 | 0.917 |
|  | 38 | 186.268 | 233.404 | 0.584 | 0.658 | 0.525 | 0.920 |
|  | 58 | 145.504 | 181.401 | 0.430 | 0.483 | 0.397 | 0.924 |
|  | 59 | 196.177 | 235.795 | 0.313 | 0.344 | 0.286 | 0.880 |
|  | 60 | 215.893 | 309.338 | 0.451 | 0.549 | 0.383 | 0.889 |
|  | 61 | 222.737 | 247.415 | 0.370 | 0.391 | 0.352 | 0.881 |
| INTERPOLATED SPLINE | 2 | 278.666 | 321.640 | 0.392 | 0.422 | 0.366 | 0.872 |
|  | 9 | 175.101 | 224.717 | 0.385 | 0.439 | 0.342 | 0.896 |
|  | 14 | 207.782 | 270.986 | 0.379 | 0.437 | 0.335 | 0.885 |
|  | 35 | 194.529 | 277.729 | 0.404 | 0.490 | 0.343 | 0.874 |
|  | 38 | 186.268 | 216.127 | 0.357 | 0.385 | 0.332 | 0.896 |
|  | 58 | 145.504 | 173.293 | 0.366 | 0.401 | 0.337 | 0.921 |
|  | 59 | 196.177 | 262.106 | 0.360 | 0.420 | 0.314 | 0.871 |
|  | 60 | 215.893 | 660.790 | 0.229 | 0.464 | 0.152 | 0.675 |
|  | 61 | 222.737 | - | - | - | - | - |
| INTERPOLATED SINC | 2 | 278.666 | 319.715 | 0.390 | 0.419 | 0.365 | 0.872 |
|  | 9 | 175.101 | 223.040 | 0.384 | 0.436 | 0.342 | 0.897 |
|  | 14 | 207.782 | 270.065 | 0.379 | 0.436 | 0.335 | 0.886 |
|  | 35 | 194.529 | 276.127 | 0.404 | 0.489 | 0.344 | 0.875 |
|  | 38 | 186.268 | 214.272 | 0.356 | 0.383 | 0.333 | 0.897 |
|  | 58 | 145.504 | 84.451 | 0.157 | 0.124 | 0.213 | 0.954 |
|  | 59 | 196.177 | 261.503 | 0.359 | 0.419 | 0.314 | 0.872 |
|  | 60 | 215.893 | 667.866 | 0.225 | 0.461 | 0.149 | 0.671 |
|  | 61 | 222.737 | 116.606 | 0.204 | 0.155 | 0.297 | 0.939 |
| INTERPOLATED TRILINEAR | 2 | 278.666 | 369.649 | 0.389 | 0.453 | 0.341 | 0.847 |
|  | 9 | 175.101 | 279.040 | 0.366 | 0.475 | 0.298 | 0.862 |
|  | 14 | 207.782 | 342.877 | 0.367 | 0.486 | 0.295 | 0.846 |
|  | 35 | 194.529 | 304.333 | 0.403 | 0.517 | 0.330 | 0.859 |
|  | 38 | 186.268 | 335.198 | 0.398 | 0.557 | 0.310 | 0.834 |
|  | 58 | 145.504 | 294.782 | 0.097 | 0.146 | 0.072 | 0.812 |
|  | 59 | 196.177 | 39.909 | 0.043 | 0.026 | 0.128 | 0.975 |
|  | 60 | 215.893 | 669.081 | 0.206 | 0.423 | 0.136 | 0.665 |
|  | 61 | 222.737 | 263.411 | 0.390 | 0.426 | 0.360 | 0.875 |

## Footnotes

2 https://nifti.nimh.nih.gov/nifti-1/

3 https://www.nitrc.org/projects/mricron

4 The software codebase will be made publicly available upon publication.

5 https://www.tensorflow.org/tensorboard

